# Maternal psychiatric diagnosis and comorbidities around pregnancy

**DOI:** 10.1101/2021.03.01.21252671

**Authors:** Vahe Khachadourian, Arad Kodesh, Stephen Z. Levine, Emma Lin, Joseph D. Buxbaum, Veerle Bergink, Sven Sandin, Abraham Reichenberg, Magdalena Janecka

**Affiliations:** Department of Psychiatry, Icahn School of Medicine at Mount Sinai, New York, New York; Seaver Autism Center for Research and Treatment, Icahn School of Medicine at Mount Sinai, New York, New York; Mindich Child Health and Development Institute, Icahn School of Medicine at Mount Sinai, New York, New York; Department of Community Mental Health, University of Haifa, Haifa, Israel; Meuhedet Health Services, Tel Aviv, Israel; Friedman Brain Institute, Icahn School of Medicine at Mount Sinai, New York, New York; Department of Obstetrics, Gynecology and Reproductive Science, Icahn School of Medicine at Mount Sinai, New York, New York; Department of Psychiatry, Erasmus MC, Rotterdam, The Netherlands; Department of Medical Epidemiology and Biostatistics, Karolinska Institutet, Stockholm, Sweden; Department of Environmental Medicine and Public Health, Icahn School of Medicine at Mount Sinai, New York, New York

## Abstract

**IMPORTANCE:** Comorbidity between mental and physical health is a frequent problem, resulting in greater morbidity than the sum of the effects of individual diseases. Despite pervasive physiological and behavioral changes during pregnancy, the pattern of comorbidities among pregnant women is not well-understood.

**OBJECTIVE:** Systematically examine the associations between mental and physical diagnoses around pregnancy.

**DESIGN, SETTING, PARTICIPANTS:** Population-based sample, containing all children born between January 1st, 1997 and December 31^st^, 2008 within a health maintenance organization (HMO) in Israel. Cohort children were linked to their mothers’ health information, creating mother-child dyads.

**MAIN OUTCOMES AND MEASURES:** The primary outcome was any ICD-9 mental health diagnosis within the pregnancy period or the preceding 12 months. The secondary outcomes were specific ICD-9 mental health diagnoses. Exposures were all ICD-9 physical (non-mental) health diagnoses received during the exposure period. We used logistic regression to test the risk of physical health diagnoses in pregnant women associated with mental health diagnoses. The covariates for this study included maternal age, birth year, socioeconomic status and total number of ICD-9 diagnoses during the exposure period.

**RESULTS:** The analytical sample included 76,783 mother-child dyads, with 30,084 unique mothers. The mean age at birth was 29.8 years, with 4.4% of mothers diagnosed with a mental health disorder during the pregnancy period. Of the 18 major ICD-9 diagnostic categories, 10 were positively associated with mental health disorders, including ICD categories of *e.g*., symptoms, signs, and ill-defined conditions (OR=1.69; 95% CI=1.50, 1.89), musculoskeletal (OR=1.33; 95% CI=1.22,1.45) and digestive system diseases (OR=1.25; 95% CI=1.15, 1.37). Comorbidity between mental and physical health diagnoses was higher than that observed between various physical health diagnoses. Associations between physical health diagnoses and specific mental health conditions were consistent with the results of the general mental health and physical health comorbidity.

**CONCLUSION AND RELEVANCE:** We observed that associations between maternal diagnoses and mental health stand out from the general pattern of comorbidity between non-mental health diseases. The high co-occurrence of mental health and physical health diagnoses has implications for diagnoses and management of morbidity among pregnant women, with potential impact on pregnancy and child health outcomes.

## INTRODUCTION

Multimorbidity, defined as co-occurrence of two or more diseases in the same person, is a major health problem affecting a substantial portion of the population^1^. Over the past decades, the prevalence of multimorbidity has been on the rise^2,3^. This increase in prevalence is partly due to the extended longevity of the baby-boom generation^1^, as well as behavioral and environmental factors fueling the growing prevalence of non-communicable diseases^4^. Comorbidity is a specific type of multimorbidity, which is defined in relation to an index disease^5,6^, i.e. the main condition under study. Comorbid health conditions have great impact on the quality of life of the patient, are costly to treat^7^ and harder to manage than single select conditions^8^. The coexistence of multiple diseases may have health effects that are greater than the sum of the effects of individual diseases^9^.

The comorbidity between mental and physical health conditions has received increasing attention over the past years^10^. Most of the studies have focused on specific pairs of health conditions, providing valuable insights about such comorbidity (e.g., depression and diabetes^11^). Furthermore, in a recent study using a population-based cohort from Denmark, Momen et al.^12^ evaluated associations between mental disorders and broad categories of medical conditions, shedding light on a wide range of potential associations between diagnoses of mental disorder and subsequent medical conditions. Nevertheless, studies systematically evaluating potential comorbidities of mental disorders with a wide spectrum of physical health conditions remain scarce, and little is still known regarding whether mental conditions are more likely to co-occur with other health conditions, compared to any other type of index disease. Furthermore, the extent to which comorbidities fluctuate over lifetime has not been examined to date, despite known effects of age on the risk of many diagnoses.

In particular, the studies exploring comorbid health conditions around pregnancy are uncommon^13^. Pregnancy is a unique period with profound physiological and behavioral changes^14^.Women with pre-pregnancy chronic medical illness require special care, because medication regimes may alter as well as the natural disease course. Moreover, pregnancy is a period associated with specific health risks for the mother due to the new onset of cardiovascular conditions, endocrine disorders or blood diseases (e.g., hypertensive disorders of pregnancy, diabetes gravidarum, and anemia^15–17^). In addition, mood and anxiety disorders are highly prevalent in women in their reproductive ages, including during the perinatal period^18,19^.

A better understanding of mental and physical health comorbidity around the pregnancy period can offer insights into diagnosis and management of maternal health conditions. Importantly, both mental and physical health problems during pregnancy have been associated with a host of adverse outcomes in offspring^15,20^ (e.g., risk of infections, asthma, obesity, cognitive performance and psychiatric disorders). There is increasing awareness that many adult disorders may have at least partly fetal origin (*e.g*. Developmental Origins of Health and Disease (DOHaD) approach^21^). Better understanding of the patterns of comorbidity in pregnancy will therefore have implications not only for maternal health, but may also shed new light on the important determinants of health outcomes for the child.

We used a large, population-based cohort to systematically and rigorously investigate the spectrum of associations between maternal mental disorders and physical health conditions just before and during pregnancy. In order to better understand unique patterns of comorbidity with mental health disorders, we also explored patterns of comorbidity among all other diagnostic categories. The overarching aim of the study was to examine the associations between mental and physical diagnoses, and put them in context by comparing them to a wide range of comorbidities between physical health conditions.

## METHODS

### Study design and population

We conducted a cohort study using a population-based sample from a large health maintenance organization (HMO) in Israel (Meuhedet), which has been described previously^22,23^. Briefly, per legislations in Israel, citizens are required to obtain medical insurance from the existing HMOs. The equivalent health plans and fee structure across HMOs, along with the regulations prohibiting HMOs from refusing a citizen membership, minimize the risk of ascertainment bias in our sample.

The cohort included all children born between January 1^st^, 1997 and December 31^st^, 2008. First, the study sample comprised 19.5% of all the births within the Meuhedet HMO during that period. Additionally, the sample included all the siblings of the children that were selected in the first stage of the sampling who were born during the same period (1997-2008).

All selected children were linked to their family records, creating mother-child dyads. Since our focus was on maternal, rather than child’s health, pregnancies leading to multiple live births (e.g., twins or triplets) were each represented by a single mother-child dyad.

To assure ascertainment of maternal diagnosis during the 12 months preceding their pregnancies, the analytical sample was restricted to pregnancies leading to a live birth between January 1^st^, 1999 through December 31^st^, 2008. All dyads where maternal age was younger than 13, or older than 55, were assumed to be due to potential administrative errors and were excluded from the sample. The study protocol was reviewed and approved by the Helsinki Ethics Committee of the Meuhedet and the institutional review board of the University of Haifa. Since the data did not include any individual identifiers, waiver of informed consent was granted by the reviewing bodies.

### Outcome

Presence or absence of psychiatric diagnosis (International Classification of Diseases, Ninth Revision (ICD-9) codes 290 through 319, including all the subcodes) during the pregnancy period and the preceding year (a total of 636 days preceding the child’s birth date) served as the main outcome variable. The maternal diagnostic codes, per ICD-9 were ascertained form the Meuhedet Diagnostic Classification Register. The hierarchical organization of diagnostic codes in ICD-9 has 4 levels, presenting information from least to most specific diagnosis (**Figure 1**). While for the primary analyses we pooled all diagnoses and only considered maternal binary status of having any mental health diagnosis during the exposure period (level 1), in additional analyses we further defined more specific mental health outcomes using the information from level 3 ICD-9 diagnostic codes with a prevalence of at least 0.1% in the sample.

**Figure 1.**
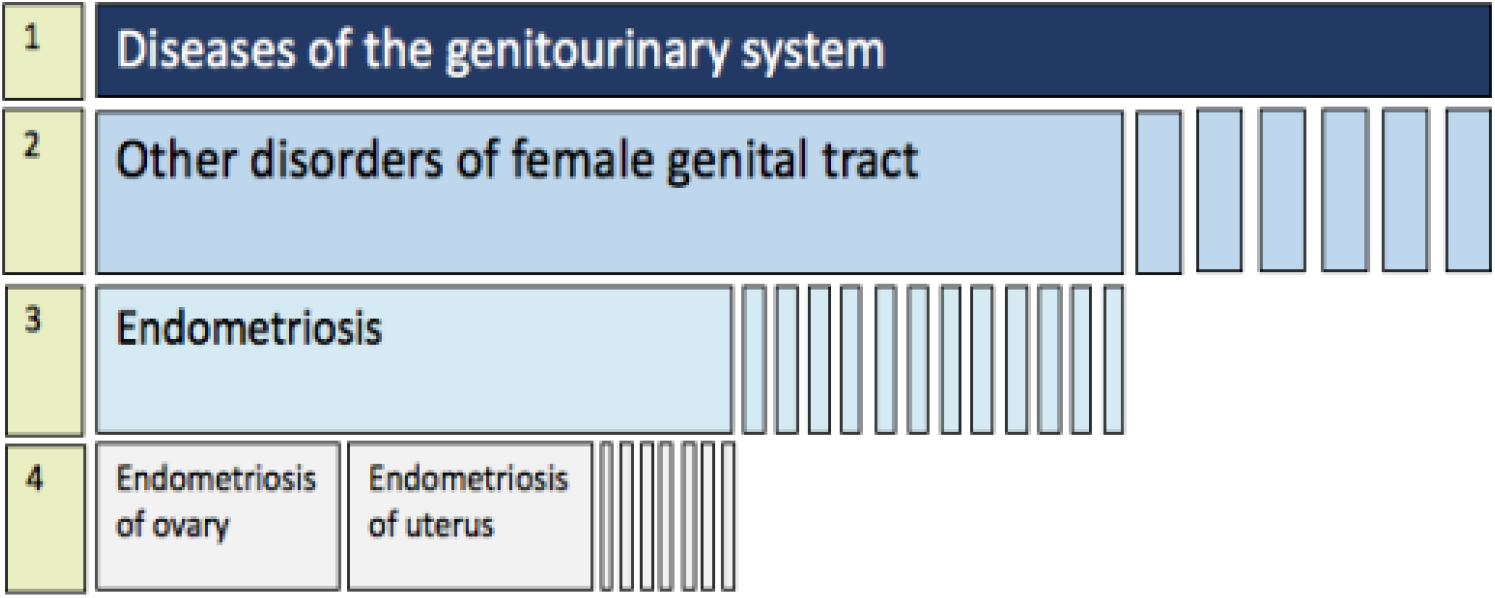
Example of the hierarchical organization of the ICD-9 taxonomy. ICD-9 categories are organized from the most general (level 1, top row), through most specific diagnostic codes (level 4, bottom row). Level 3 diagnoses were used in the current study.

### Exposure

The exposure variables in the main analyses were individual-level physical (non-mental) health diagnoses, classified at level 1 ICD-9 codes obtained from the Meuhedet Diagnostic Classification Register. In additional analyses, we further defined more specific exposure categories according to diagnostic codes at level 3, as they offer an additional level of detail about the underlying health condition. The time window for defining exposures included a total of 636 days preceding the child’s birth, i.e. including the full estimated pregnancy period (270 days), as well as the year preceding the conception. All diagnoses which were not coded according to ICD-9, and the available information did not allow for identifying and assigning them to any equivalent ICD-9, were omitted.

### Covariates

The covariates included maternal age at child’s birth, residential socioeconomic status (SES), and the total number of maternal diagnoses during the pregnancy period and the preceding year. Residential SES was a summary index based on household census data and was a function of number of electrical appliances per person and per capita income^24^. Information about socioeconomic status (SES) was obtained from Central Bureau of Statistics Registry, while the Meuhedet records served as the source for all other covariates.

### Statistical Analysis

#### Primary analysis

All analyses were performed using R programming language (version 4.0.0)^25^. The associations between maternal mental health diagnoses and other maternal ICD-9 level 1 diagnostic categories were assessed using binary logistic regression models. In order to account for potential clustering effects due to possible multiple deliveries among mothers in the study period we used clustered sandwich estimator, implemented in the *clusterSEs* package (v2.6.2)^26^.

To account for the varying risk of multiple diagnoses across the lifetime, the models included maternal age at child’s birth. The models also adjusted for SES to account for the differential likelihood of ascertaining maternal mental health^27^ and other diagnoses^28^ across the SES strata. Similarly, the total number of maternal diagnoses during the exposure period was included in the adjusted models as a proxy measure for health-seeking behaviors and healthcare utilization.

Since the sampling strategy yielded a higher probability for the inclusion of mothers with multiple deliveries, we applied inverse probability selection weighting to account for the differential selection probabilities. The weights were computed based on the number of children born to each mother during the study period. Mothers with a larger number of children received lower weights in the regression model to account for their higher probability of inclusion in the sample.

The initial analysis adjusted logistic regression models were followed by a multivariable logistic regression model that concurrently include all the level 1 diagnostic categories while also adjusting for all the other covariates (i.e. age, year of birth, SES, and total number of maternal diagnoses during the exposure window).

Log odds (ICD-9 mental health diagnosis) = β_0_ + β_1_ (ICD-9 category 1) + β_2_ (ICD-9 category 2) + … + β_n_ (ICD-9 category n) + β_n+i_ (covariate_i_)

#### Secondary analyses

We run a series of secondary analyses in order to further inform the interpretation of the results from the primary analyses, regarding associations between maternal mental and physical diagnosis (both defined as respective level 1, ICD-9 diagnostic categories).

##### General comorbidity

First, we examined the association between pairs of physical health disorders, allowing us to put the burden of comorbidity between mental and physical health (primary analysis) in the wider context of comorbidity in general. To this end, we evaluated the associations between all possible pairs of level 1 physical diagnostic categories, adjusting for covariates.

##### Comorbidity patterns across different mental health conditions

Next, we examined whether distinct mental health conditions (level 3 ICD-9) are associated with different patterns of comorbidity. To this end, we repeated the analysis as specified for the primary analyses, using as an outcome each specific level 3 ICD-9 psychiatric diagnostic category with a prevalence of >0.1% in our analytical sample. These models for specific psychiatric diagnoses included the same exposures as described for the primary analysis.

##### Specific comorbidities of mental health conditions

Finally, we investigated whether the comorbidity patterns observed in our primary analysis are likely underlain by the association between mental health disease (ICD-9 level 1) and specific physical health diagnoses (ICD-9 level 3). Given the large number of ICD-9 level 3 diagnoses, in these analyses we systematically followed a multistep approach in order to minimize potential false-positive associations. To address sparse data bias^29^ all maternal diagnoses with a recorded frequency of less than 10 in the pregnancies where mother either did, or did not receive a mental health diagnosis were excluded. Each ICD-9 level 3 diagnosis was assessed in a separate model (adjusted univariate models) that adjusted for maternal age, SES, year of birth, and total number of diagnoses received during the pregnancy period. To address potential inflation of type I error due to multiple testing, P-values were corrected for a false discovery rate (Q-value) of 5%^30^. All maternal diagnoses that remained significant after adjusting for multiple testing were jointly included into an adjusted multiple logistic regression model. **Figure S1** outlines the overview of the analytical strategy for this secondary analysis.

All secondary analyses were adjusted for the same covariates as specified in the primary analyses. Analogously, we also accounted for the presence of correlated data by computing cluster sandwich estimators^26^, and included sample weights. The robustness of our results in respect to the potential effect of missing data on study covariates were examined by sensitivity analyses comparing the results of complete case analyses with results obtained after multivariate imputation by chained equation^31^.

## RESULTS

The source population included 84,755 mother-child dyads, with children born 1999 to 2008. After removing the observations with missing values on SES (n= 7,957) and mother-child dyads with a maternal age at delivery below 13 or above 55 (n=15), the analytical sample included 76,783 mother-child dyads, including 30,085 unique maternal IDs. None of the observations had a missing value for maternal age or date of birth. Table 1 presents the analytical sample characteristics and prevalence of mental health diagnoses categories with a minimum prevalence of 0.1% (i.e. ICD-9: 293, 300, 301, 307, and 311). Women with mental health conditions had on average 26.4 physical health diagnoses recorded in the period around pregnancy, compared with 14.5 in women without any mental health conditions (P <0.001). In our sample, the burden of diagnoses within different ICD-9 level 1 categories differed between pregnancy period and periods preceding and following it (**Figure 2**; **Figure S2**).

**Table 1.** Demographic characteristics of the analytical sample (mother-child dyads)

| Variables | Any psychiatric diagnosis<br>(n = 3,353) | No psychiatric diagnosis<br>(n = 73,430) | Total<br>(N = 76,783) |
| --- | --- | --- | --- |
| Maternal age at delivery, mean (SD) | 31.4(5.2) | 29.7 (5.4) | 29.8 (5.4) |
| Socioeconomic status (SES), mean (SD) | 7.8 (4.3) | 7.6 (4.3) | 7.6 (4.3) |
| Delivery year, n (%) |  |  |  |
| 1999 | 243 (7.2%) | 7535 (10.3%) | 7778 (10.1%) |
| 2000 | 289 (8.6%) | 7565 (10.3%) | 7854 (10.2%) |
| 2001 | 290 (8.6%) | 7669 (10.4%) | 7959 (10.4%) |
| 2002 | 319 (9.5%) | 7881 (10.7%) | 8200 (10.7%) |
| 2003 | 305 (9.1%) | 8016 (10.9%) | 8321 (10.8%) |
| 2004 | 352 (10.5%) | 7858 (10.7%) | 8210 (10.7%) |
| 2005 | 434 (12.9%) | 7491 (10.2%) | 7925 (10.3%) |
| 2006 | 378 (11.3%) | 7179 (9.8%) | 7557 (9.8%) |
| 2007 | 467 (13.9%) | 7329 (10.0%) | 7796 (10.2%) |
| 2008 | 276 (8.2%) | 4907 (6.7%) | 5183 (6.8%) |
| Total number of level 3 ICD-9 diagnosis, mean (SD) | 26.4 (19.8) | 14.5 (12.4) | 15.1 (13.1) |
| Transient mental disorders due to conditions classified elsewhere (ICD9: 293); n (%) |  |  | 114 (0.1%) |
| Anxiety, dissociative and somatoform disorders (ICD9: 300); n (%) |  |  | 1571 (2.0%) |
| Personality disorders (ICD9: 301); n (%) |  |  | 78 (0.1%) |
| Mental disorder related special symptoms or syndromes not elsewhere classified (ICD9: 307); n (%) |  |  | 946 (1.2%) |
| Depressive disorder, not elsewhere classified (ICD9: 311); n (%) |  |  | 630 (0.8%) |

Note: Observations are based on pregnancies, hence, mothers with multiple pregnancies contribute multiple data points.

**Figure 2.**
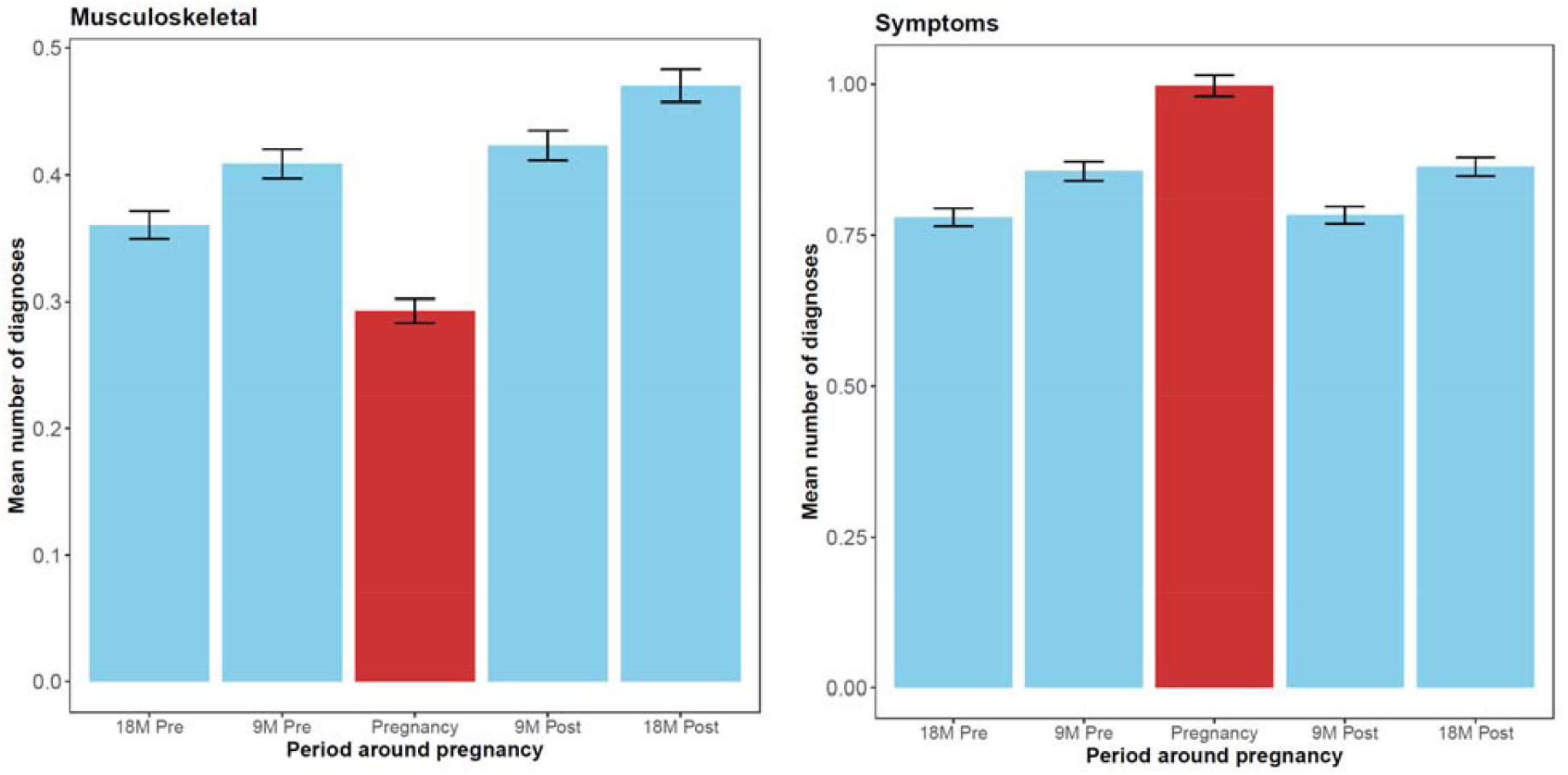
Mean number of diagnoses around pregnancy period by selected ICD-9 level 1 diagnostic categories (right: diseases of the musculoskeletal system and connective tissue, left: symptoms, signs, and ill-defined conditions)

### Associations between maternal mental health diagnosis and physical health diagnostic categories

The associations between mental and physical diagnoses (both defined at ICD-9 level 1) are presented in **Figure 3** and **Table S1**. In the logistic regression model adjusting for maternal age, SES, year of birth, total number of maternal diagnoses during the pregnancy period, and concurrently including 17 ICD-9 level 1 diagnostic categories, 10 of these level 1 diagnostic categories (59%) had a statistically significant positive association with mental health level 1 diagnosis. Maternal mental health diagnosis around pregnancy was most strongly associated with symptoms, signs, and ill-defined conditions (OR = 1.69; 95% CI = 1.50, 1.89), supplementary classification of external causes of injury and poisoning (OR = 1.39; 95% CI = 1.13, 1.71), diseases of the musculoskeletal system and connective tissue (OR = 1.33; 95% CI = 1.22,1.45).

**Figure 3.**
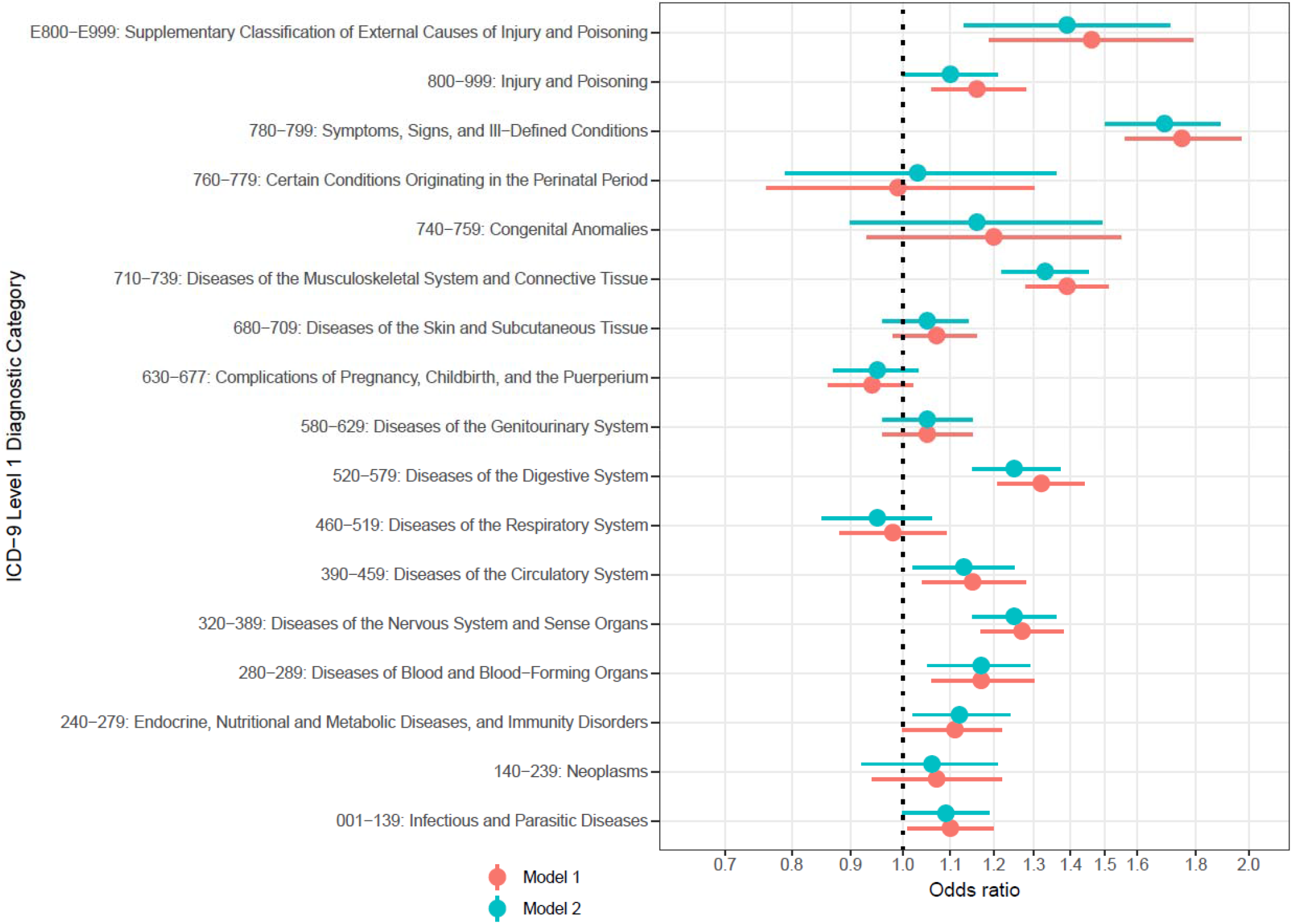
Associations between maternal ICD-9 level 1 diagnostic categories and ICD-9 level 1 mental health diagnosis Model 1 was adjusted for SES, maternal age at delivery, total number of diagnoses during 21 months period before delivery, and year of delivery Model 2 was adjusted for all variables in model one as well as all the ICD-9 level 1 diagnostic categories presented in this figure

### General comorbidity

Analyses exploring the relationship between all the possible pairs of level 1 diagnostic categories yielded several statistically significant associations (**Table 2**). Nevertheless, the average number of statistically significant associations across all other level 1 categories was 4.7, which was lower than 10.0 associations observed in the mental health outcomes. Higher comorbidity rate for mental health conditions was observed in comparison with both rare and prevalent physical health disorders, indicating that our results were not due to differential statistical power to detect associations. The average number of positive associations across all other diagnostic categories was even lower when compared with the positive associations with mental health (2.4 vs. 10.0) while the number of negative associations were higher (2.3 vs. 0.0). Some of the observed associations between the spectrum of level 1 diagnostic categories were of small magnitude, which despite being statistically significance (due to a high power) could lack practical and or clinical relevance. The median odds ratio (OR) for the positive associations between all level 1 diagnostic categories was 1.16 while for the positive associations between mental and physical health diagnoses it was 1.21.

**Table 2.** Adjusted odds ratio of the associations between ICD-9 level 1 diagnostic categories

|  |  | Outcome (ICD-9 level 1 diagnostic category) |  |  |  |  |  |  |  |  |  |  |  |  |  |  |  |  |  |
| --- | --- | --- | --- | --- | --- | --- | --- | --- | --- | --- | --- | --- | --- | --- | --- | --- | --- | --- | --- |
|  |  | MH | C17 | C16 | C15 | C14 | C13 | C12 | C11 | C10 | C9 | C8 | C7 | C6 | C5 | C4 | C3 | C2 | C1 |
| Exposure (ICD-9 level 1 diagnostic category) | C1 | 1.09* | 0.84** | 1.04 | 1.00 | 0.96 | 0.99 | 0.95* | 1.34*** | 0.89*** | 0.96 | 1.07** | 1.16*** | 1.04 | 1.04* | 0.90*** | 0.92** | 1.13*** | N/A |
|  | C2 | 1.06 | 0.94 | 1.00 | 0.89** | 0.90 | 1.64*** | 0.90** | 1.67*** | 0.80*** | 1.05 | 0.94 | 0.79*** | 1.22*** | 1.09** | 0.96 | 1.07 | N/A |  |
|  | C3 | 1.12* | 0.74*** | 0.88*** | 1.02 | 0.88 | 1.00 | 0.91*** | 0.87*** | 1.01 | 0.85*** | 0.94* | 0.74*** | 1.03 | 0.91*** | 1.27*** | N/A |  |  |
|  | C4 | 1.17** | 0.93 | 0.88*** | 1.10** | 0.94 | 0.91 | 0.94* | 1.03 | 1.23*** | 1.00 | 1.11*** | 0.86*** | 1.10** | 0.90*** | N/A |  |  |  |
|  | C5 | 1.25*** | 0.85* | 1.07** | 1.00 | 0.87* | 1.00 | 1.05* | 1.03 | 0.86*** | 0.86*** | 1.10*** | 1.10*** | 0.96 | N/A |  |  |  |  |
|  | C6 | 1.13* | 0.94 | 0.96 | 0.99 | 1.03 | 1.11 | 1.07* | 1.05 | 1.21*** | 0.87*** | 1.22*** | 0.95 | N/A |  |  |  |  |  |
|  | C7 | 0.95 | 0.77*** | 1.03 | 1.13*** | 0.73*** | 1.09 | 1.01 | 0.89*** | 0.74*** | 0.79*** | 1.11*** | N/A |  |  |  |  |  |  |
|  | C8 | 1.25*** | 0.86* | 1.02 | 1.46*** | 0.95 | 1.07 | 1.16*** | 1.02 | 0.94** | 1.01 | N/A |  |  |  |  |  |  |  |
|  | C9 | 1.05 | 0.80*** | 0.86*** | 1.09*** | 1.01 | 0.95 | 0.97 | 0.92*** | 1.07*** | N/A |  |  |  |  |  |  |  |  |
|  | C10 | 0.95 | 0.97 | 0.91*** | 0.97 | 1.46*** | 1.06 | 0.86*** | 0.85*** | N/A |  |  |  |  |  |  |  |  |  |
|  | C11 | 1.05 | 0.95 | 1.22*** | 0.88*** | 0.88 | 1.26** | 1.01 | N/A |  |  |  |  |  |  |  |  |  |  |
|  | C12 | 1.33*** | 1.82*** | 1.36*** | 1.23*** | 0.87 | 1.33*** | N/A |  |  |  |  |  |  |  |  |  |  |  |
|  | C13 | 1.16 | 0.83 | 1.17 | 0.95 | 1.00 | N/A |  |  |  |  |  |  |  |  |  |  |  |  |
|  | C14 | 1.03 | 0.57* | 1.02 | 0.99 | N/A |  |  |  |  |  |  |  |  |  |  |  |  |  |
|  | C15 | 1.69*** | 0.91 | 1.02 | N/A |  |  |  |  |  |  |  |  |  |  |  |  |  |  |
|  | C16 | 1.10* | 3.92*** | N/A |  |  |  |  |  |  |  |  |  |  |  |  |  |  |  |
|  | C17 | 1.39** | N/A |  |  |  |  |  |  |  |  |  |  |  |  |  |  |  |  |
|  | MH | N/A |  |  |  |  |  |  |  |  |  |  |  |  |  |  |  |  |  |

Note: Odds ratios are adjusted for SES, maternal age at delivery, total number of diagnoses in pregnancy, year of delivery, and all the ICD-9 level 1 diagnostic categories presented in this table; Statistically significant positive associations are highlighted in blue and negative associations are highlighted in yellow.
\* : P-value <0.05; \*\* : P-value <0.01; \*\*\* : P-value <0.001
C1: Infectious and Parasitic Diseases; C2: 140-239: Neoplasms; C3: Endocrine, Nutritional and Metabolic Diseases, and Immunity Disorders; C4: Diseases of Blood and Blood-Forming Organs; C5: Diseases of the Nervous System and Sense Organs; C6: Diseases of the Circulatory System; C7: Diseases of the Respiratory System; C8: Diseases of the Digestive System; C9: Diseases of the Genitourinary System; C10: Complications of Pregnancy, Childbirth, and the Puerperium; C11: Diseases of the Skin and Subcutaneous Tissue; C12: Diseases of the Musculoskeletal System and Connective Tissue; C13: Congenital Anomalies; C14: Certain Conditions Originating in the Perinatal Period; C15: Symptoms, Signs, and Ill-Defined Conditions; C16: Injury and Poisoning; C17: Supplementary Classification of External Causes of Injury and Poisoning; MH: Mental Disorders.

### Comorbidity patterns across different mental health conditions

The frequencies of level 1 diagnostic categories among women with and without specific psychiatric diagnoses are presented in **Table S2**. In a series of models evaluating associations of specific psychiatric diagnoses (i.e. ICD-9 level 3: 293, 300, 301, 307, 311) with different ICD-9 level 1 maternal diagnosis categories, we found a total of 15 statistically significant positive associations (**Table 3**). Different psychiatric conditions were associated with similar physical health diagnostic categories (**Table 3**). For example, ICD-9 category of Diseases of Blood and Blood-Forming Organs was associated with anxiety, dissociative and somatoform disorders (ICD-9 300; OR = 1.23; 95% CI = 1.07-1.42), and depressive disorder (ICD-9 311; OR = 1.41; 95% CI = 1.13-1.74)). For specific pairs of diagnoses that did not reach statistical significance, the direction of the effect was consistent across different psychiatric diagnoses, with strongly overlapping confidence intervals. These associations were also similar to the pattern recorded in the primary analysis (**Figure 2 and Table S1**).

**Table 3.** Associations between maternal ICD-9 level 1 diagnostic categories and specific ICD-9 level 3 psychiatric diagnoses

|  | Level 3 mental health diagnosis |  |  |  |  |
| --- | --- | --- | --- | --- | --- |
|  | Transient mental disorders due to conditions classified elsewhere ICD-9:293 | Anxiety, dissociative and somatoform disorders ICD-9:300 | Personality disorders ICD-9:301 | Mental disorder related special symptoms or syndromes not elsewhere classified ICD-9:307 | Depressive disorder, not elsewhere classified ICD-9:311 |
| Level 1 ICD-9 diagnostic category | OR (95%CI)<sup>a</sup> | OR (95%CI)<sup>a</sup> | OR (95%CI)<sup>a</sup> | OR (95%CI)<sup>a</sup> | OR (95%CI)<sup>a</sup> |
| 001-139: Infectious and Parasitic Diseases | 1.03 (0.67, 1.58) | 1.10 (0.97, 1.24) | 1.04 (0.64, 1.68) | 1.05 (0.90, 1.23) | 1.03 (0.85, 1.25) |
| 140-239: Neoplasms | 1.35 (0.67, 2.70) | 1.13 (0.94, 1.36) | 1.33 (0.56, 3.17) | 1.03 (0.80, 1.32) | 0.98 (0.73, 1.31) |
| 240-279: Endocrine, Nutritional and Metabolic Diseases, and Immunity Disorders | 1.37 (0.87, 2.17) | 1.11 (0.96, 1.27) | 1.63 (0.92, 2.90) | 1.05 (0.88, 1.27) | 1.44 (1.17, 1.76)*** |
| 280-289: Diseases of Blood and Blood-Forming Organs | 1.63 (1.03, 2.57)* | 1.23 (1.07, 1.42)** | 1.63 (0.91, 2.93) | 1.10 (0.92, 1.33) | 1.41 (1.13, 1.74)** |
| 320-389: Diseases of the Nervous System and Sense Organs | 1.22 (0.82, 1.83) | 1.12 (0.99, 1.26) | 1.35 (0.84, 2.17) | 1.89 (1.61, 2.21)*** | 1.14 (0.95, 1.38) |
| 390-459: Diseases of the Circulatory System | 1.00 (0.58, 1.73) | 1.22 (1.06, 1.41)** | 1.23 (0.66, 2.29) | 1.20 (1.01, 1.44)* | 1.21 (0.96, 1.51) |
| 460-519: Diseases of the Respiratory System | 1.10 (0.63, 1.91) | 0.98 (0.84, 1.15) | 0.58 (0.33, 1.03) | 1.03 (0.84, 1.26) | 0.92 (0.73, 1.17) |
| 520-579: Diseases of the Digestive System | 1.29 (0.87, 1.90) | 1.37 (1.21, 1.55)*** | 1.00 (0.62, 1.63) | 1.31 (1.13, 1.53)*** | 1.06 (0.88, 1.28) |
| 580-629: Diseases of the Genitourinary System | 0.74 (0.47, 1.17) | 1.09 (0.96, 1.25) | 1.46 (0.80, 2.65) | 0.99 (0.84, 1.17) | 1.07 (0.86, 1.32) |
| 630-677: Complications of Pregnancy, Childbirth, and the Puerperium | 0.79 (0.52, 1.20) | 0.99 (0.88, 1.12) | 1.51 (0.85, 2.66) | 0.90 (0.77, 1.05) | 1.00 (0.83, 1.21) |
| 680-709: Diseases of the Skin and Subcutaneous Tissue | 0.68 (0.45, 1.03) | 1.03 (0.91, 1.16) | 0.71 (0.40, 1.24) | 1.08 (0.93, 1.26) | 0.97 (0.80, 1.18) |
| 710-739: Diseases of the Musculoskeletal System and Connective Tissue | 1.59 (0.95, 2.67) | 1.36 (1.20, 1.53)*** | 0.83 (0.49, 1.39) | 1.46 (1.25, 1.70)*** | 1.00 (0.83, 1.22) |
| 740-759: Congenital Anomalies | N/A | 1.05 (0.72, 1.52) | N/A | 1.03 (0.66, 1.60) | 1.34 (0.76, 2.35) |
| 760-779: Certain Conditions Originating in the Perinatal Period | N/A | 0.91 (0.62, 1.34) | N/A | 0.92 (0.55, 1.56) | 0.90 (0.49, 1.65) |
| 780-799: Symptoms, Signs, and Ill-Defined Conditions | 2.30 (1.17, 4.52)* | 1.80 (1.50, 2.14)*** | 0.93 (0.52, 1.68) | 1.94 (1.56, 2.42)*** | 1.28 (1.00, 1.64) |
| 800-999: Injury and Poisoning | 0.81 (0.50, 1.32) | 1.05 (0.92, 1.21) | 1.39 (0.77, 2.50) | 0.99 (0.83, 1.17) | 0.97 (0.77, 1.23) |
| E800-E999: Supplementary Classification of External Causes of Injury and Poisoning | N/A | 1.05 (0.77, 1.42) | N/A | 2.26 (1.62, 3.17)*** | 0.81 (0.50, 1.34) |

<sup>a</sup> Odds ratios are adjusted for SES, maternal age at delivery, total number of diagnoses in pregnancy, year of delivery, and all the ICD-9 level 1 diagnostic categories presented in this table; Statistically significant positive associations are highlighted in blue and negative associations are highlighted in yellow.
\* :P-value <0.05; \*\* : P-value <0.01; \*\*\* : P-value <0.001

### Specific comorbidities of mental health conditions

There was a total of 786 distinct level 3 ICD-9 diagnostic codes in our sample, including 27 mental and 759 physical diagnoses. After excluding diagnostic codes recorded in <10 women with, or <10 women without, comorbid mental disorder diagnosis in pregnancy, we retained 261 diagnostic categories. In a series of logistic regression models adjusting for covariates and multiple testing, 51 of the 270 diagnoses (19%) were statistically significantly associated (48 positive associations and 3 negative associations) with receiving any mental health diagnosis (**Table S2**). **Table S2** presents all coefficient estimates with their corresponding 95% confidence intervals, Q-values (P-values adjusted for false discovery rate), and frequency of exposure diagnoses among those with and without mental health diagnosis. In a logistic regression model concurrently evaluating the associations of those 51 diagnoses with mental health diagnosis while also adjusting for the other covariates, 36 associations (14% of all eligible diagnoses, including 34 positive and 2 negative associations) remained statistically significant (**Table 4**).

**Table 4.**
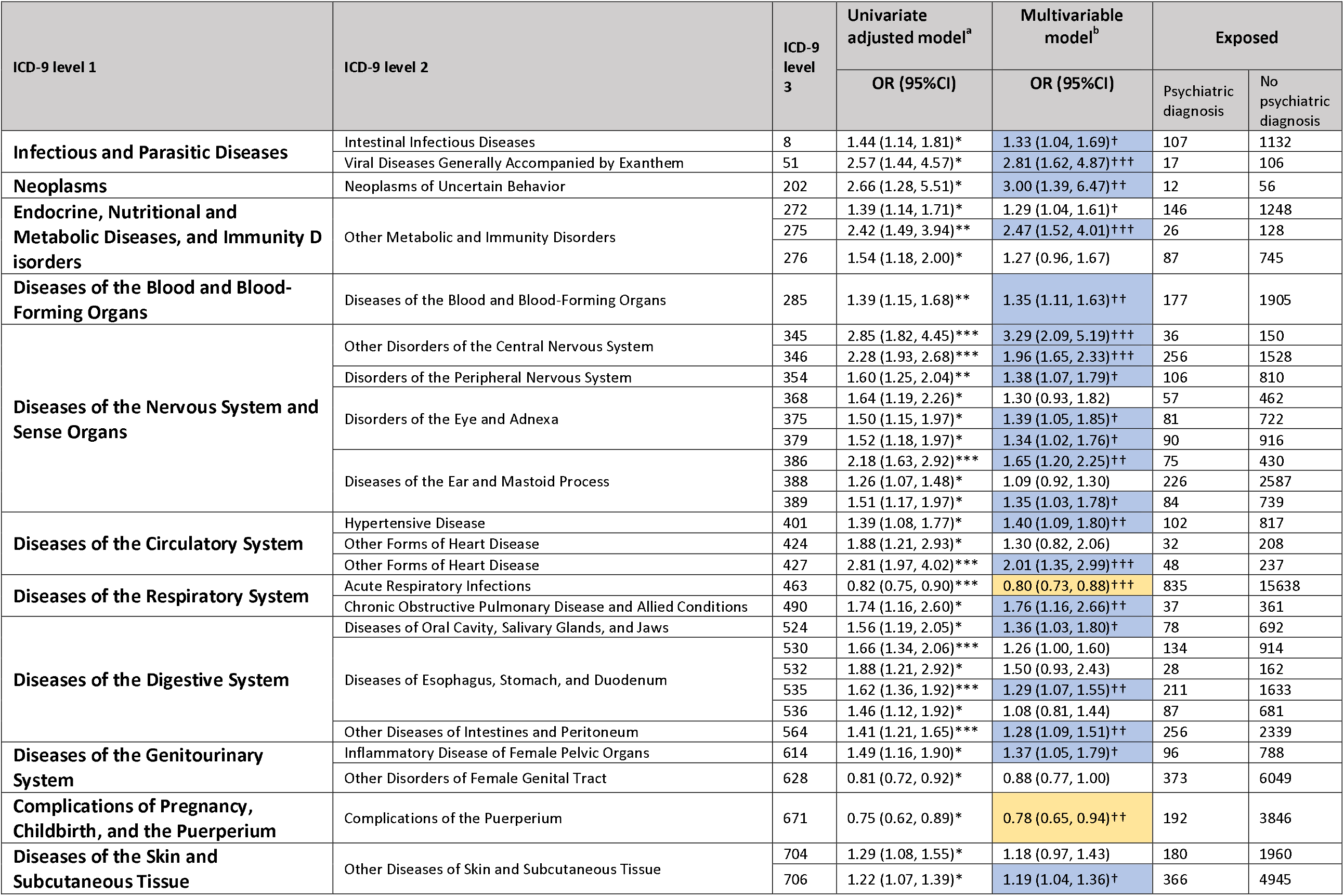
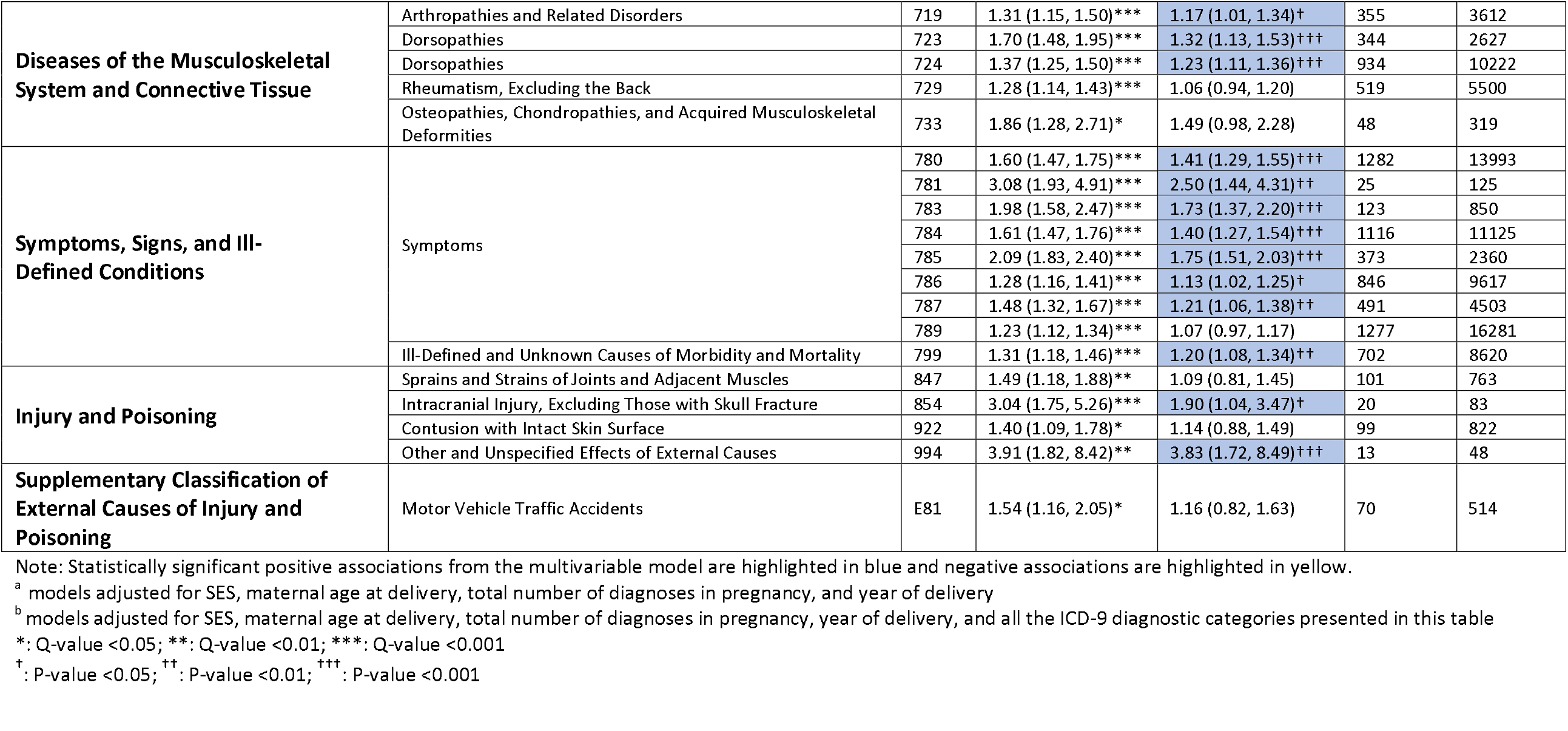
Associations between maternal ICD-9 level 3 diagnostic categories and specific ICD-9 level 1 psychiatric diagnoses

Sensitivity analysis using multiple imputation of missing variables yielded results that were near-identical to ones observed in complete case analyses, suggesting our findings were robust to the missingness pattern present in our dataset (**Table S2**).

## DISCUSSION

We found that women with a mental health disorder around pregnancy had an increased burden of disease, compared to women without any mental health diagnoses. These comorbidity rates were higher compared to pairs on physical health conditions – suggesting distinct links between mental health condition and one’s physical health around pregnancy.

This study contributes novel insights regarding the burden, and patterns of comorbidity among pregnant women. Considering that (i) pregnancy is a unique period because of physiological and behavioral changes (e.g. lower likelihood of women taking medication or consuming alcohol during pregnancy), (ii) potential occurrence of pregnancy-specific diagnoses^14^, and (iii) wide interest in investigating associations between specific maternal conditions and offspring outcomes, our results add important insights to the existing body of literature about mental health and physical health comorbidities. We highlight pervasive associations between mental and physical diagnoses, exceeding those observed between pairs of physical health conditions, with possible implications for clinical care and research. In this study we investigated the associations of major diagnostic categories, and more than 800 ICD-9 diagnoses (level 3) around pregnancy with maternal mental health diagnosis. Initial analyses identified 10 broad diagnostic categories that occur at a higher rate in women with mental health problems in the antenatal period, compared to women without such problems, including musculoskeletal, neurological, and digestive system diseases. Further analyses pointed to more specific diagnostic categories (ICD-9 level 3 diagnoses, e.g., hypertensive diseases and duodenal ulcer) significantly associated with mental health diagnoses, consistent with the existing literature about comorbidity and co-occurrence of physical and mental health conditions^32,33^. In addition to previously well-established associations, our analyses yielded indications for potentially novel or less commonly studied associations that can serve as the basis for developing new hypotheses (e.g., lacrimal system disorders, and disorders of fluid, electrolyte, and acid-base balance). Drawing the methodological approaches from our previous work^34^ allowed for direct comparison between these methodologically homogenously estimated associations with respect to direction, strength, and precision, both within this study and with previous studies focusing of the topic.

Importantly, we showed that there was no support in our data for the hypothesis that the patterns of physical comorbidities may vary between different mental health conditions. The proportion and pattern of ICD-9 level 1 diagnostic categories associated with specific mental health diagnoses were relatively similar with the exception of ICD-9 301 (personality disorders). Although the measures of associations between physical diseases and personality disorders were not statistically significant (at least partly due to low prevalence of personality disorders in the study sample), their direction and magnitude were consistent with the direction and magnitude of the observed associations between physical diseases and other specific mental health disorders.

The study findings have several implications. The wide range of diagnoses evaluated for their associations with the presence of any mental health diagnoses in general, as well as with specific psychiatric diagnoses, can offer clinical practitioners’ further insights regarding risk stratification and screening of pregnant women for comorbid mental health conditions. The study results confirm the need for screening for mental disorders during pregnancy, a practice recommended by the obstetric guidelines^35^. Clinicians should also be specifically aware of the high co-occurrence of physical diseases (e.g., neurological, gastrointestinal, and musculoskeletal diseases) among patients with mental health diagnosis. The identification of comorbid conditions is of great importance as comorbidities can worsen the course of disease for all the diseases that are present^9,36^, therefore, availability of information about risk and or existence of comorbidities might also influence treatment and management approaches.

For researchers, this study can offer insights into the complex network of associations between maternal health conditions and their impact on pregnancy and child outcomes. From the methodical perspective, our results can also inform selection of confounding variables in studies assessing impact of certain maternal diagnoses on health outcomes in the offspring. Even in situations when data on these measures are not available, pervasive nature of comorbidity should be acknowledged when drawing inferences from associations between maternal health and offspring outcomes.

The rigor and systematic nature of our approach should not be inferred as an evidence for causality between maternal diagnoses and psychiatric diagnosis. Although overall our findings are consistent with the existing literature about high comorbidity between mental and physical health outcomes^32,33^, the underlying mechanisms of the observed associations in our sample remain to be explored. While the presence of a mental health condition could cause physical health outcomes (and vice-versa), shared underlying mechanisms should also be considered.

These different mechanisms that could underlie the observed patterns of comorbidity are not mutually exclusive. Teasing out causal relationship will require careful consideration of confounding variables, temporality between the exposure and outcome of interest, and experimental work.

While this study represents a novel contribution to investigating comorbidities between mental and physical diagnoses, its limitations should be clearly acknowledged. Since the exposure window was defined as pregnancy period and one year preceding it, the chronic conditions, particularly those adequately managed without requiring frequent health care visits, were less likely to be ascertained. Although we had adequate power to analyze associations between several diagnoses and psychiatric diagnoses, there were less-common diagnoses that were excluded due to having a frequency of less than 10 across the outcome categories. In the light of these limitations, the identified associations in this study should not be considered exhaustive. Although defining the exposure based on at least 1 record of the specific diagnosis codes offered a higher sensitivity for identifying maternal diagnoses, it might have affected our specificity, leading to false positive associations (e.g. due to administrative errors, or misdiagnosis). The study had limited data for covariate control. Lack of data on routinely adjusted health behaviors, including smoking, alcohol use, and physical activity did not allow us to investigate the possible behavioral factors that could lead to the observed associations between certain pairs of disorders. Additionally, the socioeconomic status measure in our dataset was based on residential socioeconomic status and it did not include household or individual level information. Although, we adjusted for residential socioeconomic status, it could not capture the full depth of individual and household socioeconomic status ^37,38^, therefore the socioeconomic status adjustment in our analyses should be deemed incomplete. Finally, the analyses adjusted for the total number of diagnoses during the exposure period, which to certain extent can account for differential health care access and utilization. Nevertheless, lack of accurate direct measurements of health care utilization in our study remains a limitation.

In conclusion, this study followed a novel approach, supplementing prior research^34^ with new insights on associations between a wide range of maternal diagnoses and mental health diagnoses around pregnancy. We demonstrated the high burden of physical comorbidities among pregnant women with an ICD-9 diagnosis of mental health condition, especially symptoms, signs, and ill-defined conditions as well as musculoskeletal, neurological, and respiratory diseases. The degree of comorbidity was higher than for other pairs on physical health conditions, suggesting unique relationship between mental health problems and physical health. The study findings draw attention to the importance of diagnosis and management of comorbidity during pregnancy period. Considering the specific focus of our study on the period around pregnancy, any improvement in detection and management of comorbid conditions could also impact pregnancy and child health outcomes.

## Data Availability

Data access rules do not permit public sharing of the data. Interested researchers should discuss access options with Arad Kodesh and Stephen Levine.

## ACKNOWLEDGEMENTS

We would like to acknowledge the generous support of the Seaver Foundation. VK was supported by National Institute of Mental Health Award T32 MH122394. The content is solely the responsibility of the authors and does not necessarily represent the official views of the NIH or the authors’ employers.

## Notes

### Competing Interest Statement

The authors have declared no competing interest.

### Author Declarations

The study protocol was reviewed and approved by the Helsinki Ethics Committee of the Meuhedet and the institutional review board of the University of Haifa. Since the data did not include any individual identifiers, waiver of informed consent was granted by the reviewing bodies.

## REFERENCES

1. Salive ME. Multimorbidity in Older Adults. Epidemiol Rev. 2013;35(1):75–83. DOI:10.1093/epirev/mxs009

2. van den Akker M, Vaes B, Goderis G, Van Pottelbergh G, De Burghgraeve T, Henrard S. Trends in multimorbidity and polypharmacy in the Flemish-Belgian population between 2000 and 2015. Devleesschauwer B, ed. PLoS One. 2019;14(2):e0212046. DOI:10.1371/journal.pone.0212046

3. King DE, Xiang J, Pilkerton CS. Multimorbidity Trends in United States Adults, 1988–2014. J Am Board Fam Med. 2018;31(4):503–513. DOI:10.3122/jabfm.2018.04.180008

4. Singer L, Green M, Rowe F, Ben-Shlomo Y, Morrissey K. Social determinants of multimorbidity and multiple functional limitations among the ageing population of England, 2002–2015. SSM - Popul Heal. 2019;8:100413. DOI:10.1016/j.ssmph.2019.100413

5. Feinstein AR. The pre-therapeutic classification of co-morbidity in chronic disease. J Chronic Dis. 1970;23(7):455–468. DOI:10.1016/0021-9681(70)90054-8

6. Schellevis FG, van der Velden J, van de Lisdonk E, van Eijk JT, van Weel C. Comorbidity of chronic diseases in general practice. J Clin Epidemiol. 1993;46(5):469–473. DOI:10.1016/0895-4356(93)90024-u

7. Cortaredona S, Ventelou B. The extra cost of comorbidity: multiple illnesses and the economic burden of non-communicable diseases. BMC Med. 2017;15(1):216. DOI:10.1186/s12916-017-0978-2

8. Vogeli C, Shields AE, Lee TA, et al. Multiple Chronic Conditions: Prevalence, Health Consequences, and Implications for Quality, Care Management, and Costs. J Gen Intern Med. 2007;22(S3):391–395. DOI:10.1007/s11606-007-0322-1

9. Leucht S, Burkard T, Henderson J, Maj M, Sartorius N. Physical illness and schizophrenia: a review of the literature. Acta Psychiatr Scand. 2007;116(5):317–333. DOI:10.1111/j.1600-0447.2007.01095.x

10. DE Hert M, Correll CU, Bobes J, et al. Physical illness in patients with severe mental disorders. I. Prevalence, impact of medications and disparities in health care. World Psychiatry. 2011;10(1):52–77. DOI:10.1002/j.2051-5545.2011.tb00014.x

11. Lustman PJ, Griffith LS, Gavard JA, Clouse RE. Depression in adults with diabetes. Diabetes Care. Published online 1992. DOI:10.2337/diacare.15.11.1631

12. Momen NC, Plana-Ripoll O, Agerbo E, et al. Association between Mental Disorders and Subsequent Medical Conditions. N Engl J Med. 2020;382(18):1721–1731. DOI:10.1056/NEJMoa1915784

13. Reiter SF, Veiby G, Daltveit AK, Engelsen BA, Gilhus NE. Psychiatric comorbidity and social aspects in pregnant women with epilepsy - The Norwegian Mother and Child Cohort Study. Epilepsy Behav. Published online 2013. DOI:10.1016/j.yebeh.2013.08.016

14. Moya J, Phillips L, Sanford J, Wooton M, Gregg A, Schuda L. A review of physiological and behavioral changes during pregnancy and lactation: Potential exposure factors and data gaps. J Expo Sci Environ Epidemiol. Published online 2014. DOI:10.1038/jes.2013.92

15. Bansil P, Kuklina E V., Meikle SF, et al. Maternal and Fetal Outcomes Among Women with Depression. J Women’s Heal. 2010;19(2):329–334. DOI:10.1089/jwh.2009.1387

16. Horowitz KM, Ingardia CJ, Borgida AF. Anemia in Pregnancy. Clin Lab Med. Published online 2013. DOI:10.1016/j.cll.2013.03.016

17. Hernández-Díaz S, Toh S, Cnattingius S. Risk of pre-eclampsia in first and subsequent pregnancies: prospective cohort study. BMJ. 2009;338:b2255. DOI:10.1136/bmj.b2255

18. Kessler RC, Berglund P, Demler O, Jin R, Merikangas KR, Walters EE. Lifetime prevalence and age-of-onset distributions of DSM-IV disorders in the national comorbidity survey replication. Arch Gen Psychiatry. Published online 2005. DOI:10.1001/archpsyc.62.6.593

19. Vesga-López O, Blanco C, Keyes K, Olfson M, Grant BF, Hasin DS. Psychiatric disorders in pregnant and postpartum women in the United States. Arch Gen Psychiatry. 2008;65(7):805–815. DOI:10.1001/archpsyc.65.7.805

20. Ng AT, Duan L, Win T, Spencer HT, Lee M-S. Maternal and fetal outcomes in pregnant women with heart failure. Heart. 2018;104(23):1949–1954. DOI:10.1136/heartjnl-2018-313156

21. Wadhwa PD, Buss C, Entringer S, Swanson JM. Developmental origins of health and disease: brief history of the approach and current focus on epigenetic mechanisms. Semin Reprod Med. 2009;27(5):358–368. DOI:10.1055/s-0029-1237424

22. Janecka M, Kodesh A, Levine SZ, et al. Association of Autism Spectrum Disorder With Prenatal Exposure to Medication Affecting Neurotransmitter Systems. JAMA Psychiatry. 2018;75(12):1217. DOI:10.1001/jamapsychiatry.2018.2728

23. Levine SZ, Kodesh A, Viktorin A, et al. Association of Maternal Use of Folic Acid and Multivitamin Supplements in the Periods Before and During Pregnancy With the Risk of Autism Spectrum Disorder in Offspring. JAMA Psychiatry. 2018;75(2):176. DOI:10.1001/jamapsychiatry.2017.4050

24. Central Bureau of Statistics. Demographic Characteristics of the Population in Localities and Statistical Areas Jerusalem, Israel: Central Bureau of Statistics.; 1995.

25. Team RC. R: A Language and Environment for Statistical Computing. R Found Stat Comput. Published online 2016.

26. Cameron AC, Gelbach JB, Miller DL. Robust Inference With Multiway Clustering. J Bus Econ Stat. 2011;29(2):238–249. DOI:10.1198/jbes.2010.07136

27. Hudson CG. Socioeconomic status and mental illness: Tests of the social causation and selection hypotheses. Am J Orthopsychiatry. Published online 2005. DOI:10.1037/0002-9432.75.1.3

28. Dalstra JAA, Kunst AE, Borell C, et al. Socioeconomic differences in the prevalence of common chronic diseases: An overview of eight European countries. Int J Epidemiol. Published online 2005. DOI:10.1093/ije/dyh386

29. Greenland S, Mansournia MA, Altman DG. Sparse data bias: a problem hiding in plain sight. BMJ. 2016;352:i1981. DOI:10.1136/bmj.i1981

30. Benjamini Y, Hochberg Y. Controlling the False Discovery Rate: A Practical and Powerful Approach to Multiple Testing. J R Stat Soc Ser B. Published online 1995. DOI:10.1111/j.2517-6161.1995.tb02031.x

31. Zhang Z. Multiple imputation with multivariate imputation by chained equation (MICE) package. Ann Transl Med. Published online 2016. DOI:10.3978/j.issn.2305-5839.2015.12.63

32. Stein DJ, Aguilar-Gaxiola S, Alonso J, et al. Associations between mental disorders and subsequent onset of hypertension. Gen Hosp Psychiatry. 2014;36(2):142–149. DOI:10.1016/j.genhosppsych.2013.11.002

33. Lee YB, Yu J, Choi HH, et al. The association between peptic ulcer diseases and mental health problems: A population-based study: a STROBE compliant article. Medicine (Baltimore). 2017;96(34):e7828. DOI:10.1097/MD.0000000000007828

34. Kodesh A, Levine SZ, Khachadourian V, et al. Maternal health around pregnancy and autism risk: a population-based study. medRxiv. Published online January 1, 2020:2020.05.19.20089581. DOI:10.1101/2020.05.19.20089581

35. Kendig S, Keats JP, Hoffman MC, et al. Consensus Bundle on Maternal Mental Health: Perinatal Depression and Anxiety. Obstet Gynecol. 2017;129(3):422–430. DOI:10.1097/AOG.0000000000001902

36. Sartorious N. Comorbidity of mental and physical diseases: a main challenge for medicine of the 21st century. Shanghai Arch Psychiatry. 2013;25(2):68–69. DOI:10.3969/j.issn.1002-0829.2013.02.002

37. Demissie K, Hanley JA, Menzies D, Joseph L, Ernst P. Agreement in measuring socio-economic status: Area-based versus individual measures. Chronic Dis Can. Published online 2000.

38. Shavers VL. Measurement of socioeconomic status in health disparities research. J Natl Med Assoc. Published online 2007. DOI:10.13016/avw3-9cvx

