## Supplemental materials for "Maternal psychiatric diagnosis and comorbidities around pregnancy"

**SUPPLEMENTARY MATERIALS**

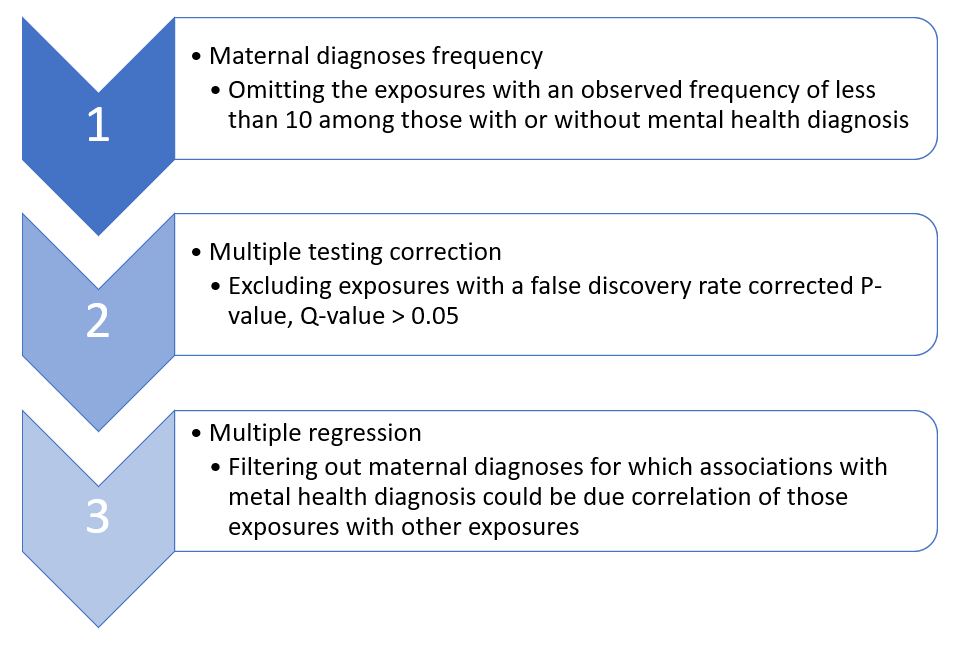

**Figure S1.** Outline of analytical steps for filtering out potential false positive associations between maternal diagnoses and mental health diagnosis

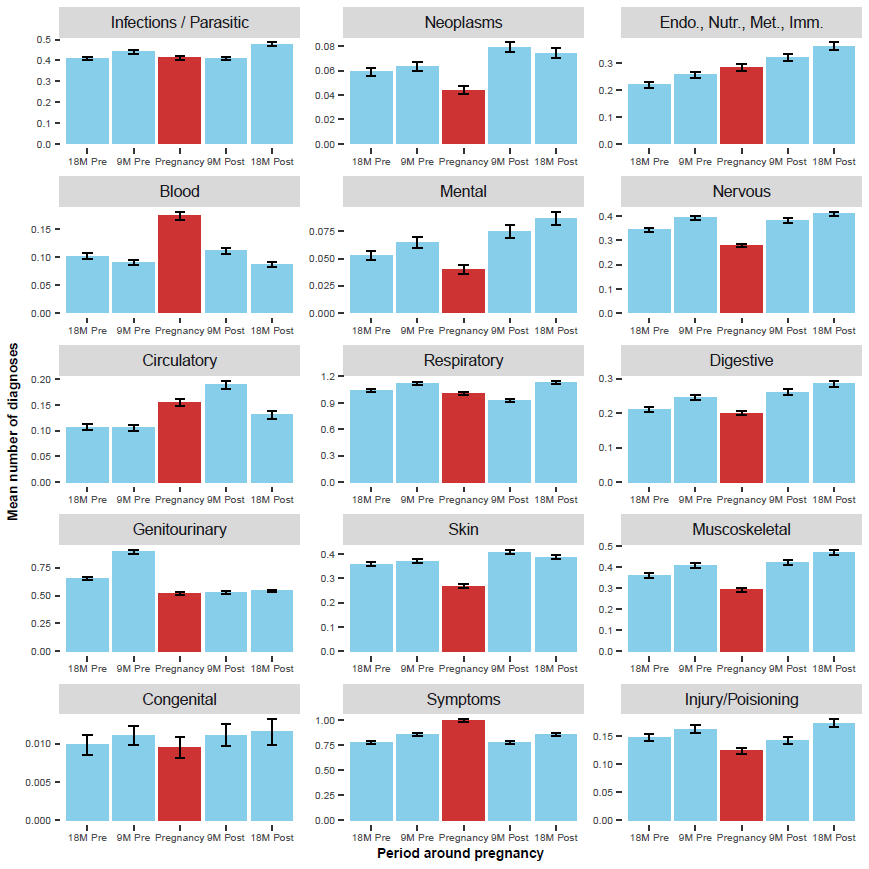

**Figure S2.** Mean number of diagnoses around pregnancy period by selected ICD-9 level 1 diagnostic categories

**Table S1.** Associations between maternal ICD-9 level 1 diagnostic categories and ICD-9 level 1 mental health diagnosis

|  | **Univariate adjusted logistic regression model^a^** | **Multivariable logistic regression model^b^** | **Exposed** | | | |
| --- | --- | --- | --- | --- | --- | --- |
|  |  |  | **Any mental health diagnosis (ICD-19: 290-319), N= 3353** | | **No mental health diagnosis, N= 73423** | |
| **Level 1 ICD-9 diagnostic category** | **OR (95%CI)** | **OR (95%CI)** | **n** | **%** | **n** | **%** |
| **001-139:** Infectious and Parasitic Diseases | 1.10 (1.01, 1.20)* | 1.09 (1.00, 1.19)† | 1910 | 57.0% | 30581 | 41.7% |
| **140-239:** Neoplasms | 1.07 (0.94, 1.22) | 1.06 (0.92, 1.21) | 358 | 10.7% | 4820 | 6.6% |
| **240-279:** Endocrine, Nutritional and Metabolic Diseases, and Immunity Disorders | 1.11 (1.00, 1.22) | 1.12 (1.02, 1.24)† | 913 | 27.2% | 11132 | 15.2% |
| **280-289:** Diseases of Blood and Blood-Forming Organs | 1.17 (1.06, 1.30)** | 1.17 (1.05, 1.29)†† | 754 | 22.5% | 10030 | 13.7% |
| **320-389:** Diseases of the Nervous System and Sense Organs | 1.27 (1.17, 1.38)*** | 1.25 (1.15, 1.36)††† | 1701 | 50.7% | 24292 | 33.1% |
| **390-459:** Diseases of the Circulatory System | 1.15 (1.04, 1.28)* | 1.13 (1.02, 1.25)† | 741 | 22.1% | 9651 | 13.1% |
| **460-519:** Diseases of the Respiratory System | 0.98 (0.88, 1.09) | 0.95 (0.85, 1.06) | 2662 | 79.4% | 47637 | 64.9% |
| **520-579:** Diseases of the Digestive System | 1.32 (1.21, 1.44)*** | 1.25 (1.15, 1.37)††† | 1365 | 40.7% | 16732 | 22.8% |
| **580-629:** Diseases of the Genitourinary System | 1.05 (0.96, 1.15) | 1.05 (0.96, 1.15) | 2177 | 64.9% | 35058 | 47.7% |
| **630-677:** Complications of Pregnancy, Childbirth, and the Puerperium | 0.94 (0.86, 1.02) | 0.95 (0.87, 1.03) | 1726 | 51.5% | 28772 | 39.2% |
| **680-709:** Diseases of the Skin and Subcutaneous Tissue | 1.07 (0.98, 1.16) | 1.05 (0.96, 1.14) | 1548 | 46.2% | 24274 | 33.1% |
| **710-739:** Diseases of the Musculoskeletal System and Connective Tissue | 1.39 (1.28, 1.51)*** | 1.33 (1.22, 1.45)††† | 1669 | 49.8% | 20668 | 28.1% |
| **740-759:** Congenital Anomalies | 1.20 (0.93, 1.55) | 1.16 (0.90, 1.49) | 87 | 2.6% | 1038 | 1.4% |
| **760-779:** Certain Conditions Originating in the Perinatal Period | 0.99 (0.76, 1.30) | 1.03 (0.79, 1.36) | 73 | 2.2% | 1317 | 1.8% |
| **780-799:** Symptoms, Signs, and Ill-Defined Conditions | 1.75 (1.56, 1.97)*** | 1.69 (1.50, 1.89)††† | 2770 | 82.6% | 41629 | 56.7% |
| **800-999:** Injury and Poisoning | 1.16 (1.06, 1.28)** | 1.10 (1.00, 1.21)† | 907 | 27.1% | 12041 | 16.4% |
| **E800-E999:** Supplementary Classification of External Causes of Injury and Poisoning | 1.46 (1.19, 1.79)*** | 1.39 (1.13, 1.71)†† | 143 | 4.3% | 1291 | 1.8% |
| ^a^ models adjusted for SES, maternal age at delivery, total number of diagnoses in pregnancy, year of delivery  ^b^ model adjusted for SES, maternal age at delivery, total number of diagnoses during 21 months period before delivery, year of delivery, and all the ICD-9 level 1 diagnostic categories presented in this table  * : P-value <0.05; ** : P-value <0.01; *** :P-value <0.001 | | | | | | |

**Table S2.** Frequencies of exposure to ICD-9 level 1 diagnostic categories by specific psychiatric diagnoses status

| **Level 1 ICD-9 diagnostic category** | **ICD-9:293** | | **ICD-9:300** | | **ICD-9:301** | | **ICD-9:307** | | **ICD-9:311** | |
| --- | --- | --- | --- | --- | --- | --- | --- | --- | --- | --- |
|  | **Yes** | **No** | **Yes** | **No** | **Yes** | **No** | **Yes** | **No** | **Yes** | **No** |
| **001-139:** Infectious and Parasitic Diseases | 66 | 32425 | 888 | 31603 | 43 | 32448 | 546 | 31945 | 356 | 32135 |
| **140-239:** Neoplasms | 11 | 5167 | 172 | 5006 | 10 | 5168 | 94 | 5084 | 71 | 5107 |
| **240-279:** Endocrine, Nutritional and Metabolic Diseases, and Immunity Disorders | 41 | 12004 | 446 | 11599 | 26 | 12019 | 222 | 11823 | 220 | 11825 |
| **280-289:** Diseases of Blood and Blood-Forming Organs | 37 | 10747 | 376 | 10408 | 21 | 10763 | 207 | 10577 | 161 | 10623 |
| **320-389:** Diseases of the Nervous System and Sense Organs | 60 | 25933 | 777 | 25216 | 38 | 25955 | 572 | 25421 | 302 | 25691 |
| **390-459:** Diseases of the Circulatory System | 27 | 10365 | 386 | 10006 | 19 | 10373 | 214 | 10178 | 150 | 10242 |
| **460-519:** Diseases of the Respiratory System | 93 | 50206 | 1255 | 49044 | 57 | 50242 | 768 | 49531 | 488 | 49811 |
| **520-579:** Diseases of the Digestive System | 53 | 18044 | 686 | 17411 | 29 | 18068 | 398 | 17699 | 242 | 17855 |
| **580-629:** Diseases of the Genitourinary System | 68 | 37167 | 1038 | 36197 | 57 | 37178 | 612 | 36623 | 405 | 36830 |
| **630-677:** Complications of Pregnancy, Childbirth, and the Puerperium | 60 | 30438 | 842 | 29656 | 50 | 30448 | 459 | 30039 | 347 | 30151 |
| **680-709:** Diseases of the Skin and Subcutaneous Tissue | 46 | 25776 | 736 | 25086 | 27 | 25795 | 444 | 25378 | 282 | 25540 |
| **710-739:** Diseases of the Musculoskeletal System and Connective Tissue | 64 | 22273 | 810 | 21527 | 32 | 22305 | 500 | 21837 | 274 | 22063 |
| **740-759:** Congenital Anomalies | 2 | 1123 | 36 | 1089 | 3 | 1122 | 26 | 1099 | 17 | 1108 |
| **760-779:** Certain Conditions Originating in the Perinatal Period | 1 | 1389 | 32 | 1358 | 1 | 1389 | 21 | 1369 | 12 | 1378 |
| **780-799:** Symptoms, Signs, and Ill-Defined Conditions | 103 | 44296 | 1329 | 43070 | 63 | 44336 | 797 | 43602 | 510 | 43889 |
| **800-999:** Injury and Poisoning | 26 | 12922 | 424 | 12524 | 27 | 12921 | 250 | 12698 | 145 | 12803 |
| **E800-E999:** Supplementary Classification of External Causes of Injury and Poisoning | 7 | 1427 | 57 | 1377 | 6 | 1428 | 50 | 1384 | 20 | 1414 |

Note: Cells with a frequency of less than 10 are highlighted in yellow.

**Table S3.** Associations between maternal ICD-9 level 3 diagnostic categories and specific ICD-9 level 1 psychiatric diagnoses

| **ICD-9** | **Diagnosis / Medical code** | **OR (95%CI)^a^** | **Exposed** | |
| --- | --- | --- | --- | --- |
|  |  |  | Psychiatric diagnosis | Psychiatric diagnosis |
| 8 | Intestinal infections due to other organisms | 1.44 (1.14, 1.81)* | 107 | 1132 |
| 9 | Ill-defined intestinal infections | 1.13 (0.94, 1.35) | 178 | 2462 |
| 34 | Streptococcal sore throat and scarlet fever | 0.91 (0.78, 1.06) | 261 | 4240 |
| 51 | Cowpox and paravaccinia | 2.57 (1.44, 4.57)* | 17 | 106 |
| 53 | Herpes zoster | 1.18 (0.72, 1.95) | 21 | 225 |
| 54 | Herpes simplex | 1.18 (0.97, 1.43) | 160 | 2021 |
| 57 | Other viral exanthemata | 1.08 (0.56, 2.09) | 12 | 187 |
| 70 | Viral hepatitis | 1.27 (0.74, 2.17) | 16 | 216 |
| 75 | Infectious mononucleosis | 0.97 (0.67, 1.42) | 39 | 496 |
| 77 | Other diseases of conjunctiva due to viruses and Chlamydiae | 1.02 (0.70, 1.48) | 42 | 809 |
| 78 | Other diseases due to viruses and Chlamydiae | 1.08 (0.93, 1.25) | 262 | 4227 |
| 79 | Viral and chlamydial infection in conditions classified elsewhere and of unspecified site | 0.98 (0.88, 1.09) | 619 | 9422 |
| 110 | Dermatophytosis | 1.03 (0.91, 1.17) | 412 | 6288 |
| 111 | Dermatomycosis, other and unspecified | 1.21 (0.94, 1.55) | 92 | 1208 |
| 112 | Candidiasis | 1.11 (1.00, 1.24) | 554 | 7789 |
| 127 | Other intestinal helminthiases | 1.25 (0.92, 1.70) | 57 | 654 |
| 128 | Other and unspecified helminthiases | 0.82 (0.49, 1.38) | 21 | 364 |
| 132 | Pediculosis and phthirus infestation | 0.86 (0.44, 1.68) | 12 | 145 |
| 133 | Acariasis | 0.60 (0.34, 1.07) | 15 | 285 |
| 202 | Other malignant neoplasms of lymphoid and histiocytic tissue | 2.66 (1.28, 5.51)* | 11 | 68 |
| 214 | Lipoma | 1.53 (1.00, 2.34) | 33 | 305 |
| 216 | Benign neoplasm of skin | 0.98 (0.84, 1.15) | 248 | 3667 |
| 218 | Uterine leiomyoma | 0.83 (0.49, 1.40) | 17 | 203 |
| 228 | Hemangioma and lymphangioma, any site | 1.01 (0.59, 1.73) | 18 | 193 |
| 238 | Neoplasm of uncertain behavior of other and unspecified sites and tissues | 2.45 (1.19, 5.07) | 12 | 56 |
| 240 | Simple and unspecified goiter | 1.13 (0.73, 1.74) | 30 | 312 |
| 241 | Nontoxic nodular goiter | 1.07 (0.65, 1.76) | 23 | 257 |
| 242 | Thyrotoxicosis with or without goiter | 0.92 (0.63, 1.34) | 43 | 589 |
| 244 | Acquired hypothyroidism | 0.81 (0.66, 0.99) | 155 | 2205 |
| 245 | Thyroiditis | 0.82 (0.55, 1.23) | 36 | 482 |
| 246 | Other disorders of thyroid | 1.04 (0.72, 1.51) | 42 | 460 |
| 250 | Diabetes mellitus | 1.03 (0.77, 1.37) | 68 | 732 |
| 253 | Disorders of the pituitary gland and its hypothalamic control | 1.27 (0.89, 1.82) | 40 | 427 |
| 256 | Ovarian dysfunction | 1.18 (0.86, 1.62) | 53 | 573 |
| 263 | Other and unspecified protein-calorie malnutrition | 1.32 (0.62, 2.78) | 13 | 142 |
| 266 | Deficiency of B-complex components | 1.13 (0.96, 1.32) | 244 | 2900 |
| 268 | Vitamin D deficiency | 1.18 (0.61, 2.28) | 13 | 107 |
| 269 | Other nutritional deficiencies | 1.12 (0.79, 1.60) | 48 | 560 |
| 272 | Disorders of lipoid metabolism | 1.39 (1.14, 1.71)* | 146 | 1248 |
| 275 | Disorders of mineral metabolism | 2.42 (1.49, 3.94)** | 26 | 128 |
| 276 | Disorders of fluid, electrolyte, and acid-base balance | 1.54 (1.18, 2.00)* | 87 | 745 |
| 277 | Other and unspecified disorders of metabolism | 0.93 (0.51, 1.70) | 14 | 123 |
| 278 | Overweight, obesity and other hyperalimentation | 1.23 (1.03, 1.47) | 206 | 1959 |
| 280 | Iron deficiency anemias | 1.12 (1.00, 1.26) | 491 | 6700 |
| 281 | Other deficiency anemias | 1.06 (0.77, 1.44) | 63 | 747 |
| 282 | Hereditary hemolytic anemias | 0.59 (0.32, 1.07) | 14 | 261 |
| 285 | Other and unspecified anemias | 1.39 (1.15, 1.68)** | 177 | 1905 |
| 286 | Coagulation defects | 1.12 (0.72, 1.75) | 26 | 290 |
| 287 | Purpura and other hemorrhagic conditions | 1.03 (0.70, 1.51) | 38 | 591 |
| 288 | Diseases of white blood cells | 1.24 (0.72, 2.14) | 21 | 238 |
| 289 | Other diseases of blood and blood-forming organs | 0.88 (0.62, 1.24) | 47 | 647 |
| 337 | Disorders of the autonomic nervous system | 1.49 (0.84, 2.64) | 19 | 123 |
| 345 | Epilepsy and recurrent seizures | 2.85 (1.82, 4.45)*** | 36 | 150 |
| 346 | Migraine | 2.28 (1.93, 2.68)*** | 256 | 1528 |
| 350 | Trigeminal nerve disorders | 1.66 (0.76, 3.61) | 10 | 71 |
| 351 | Facial nerve disorders | 1.07 (0.50, 2.26) | 11 | 105 |
| 353 | Nerve root and plexus disorders | 1.38 (0.76, 2.51) | 17 | 128 |
| 354 | Mononeuritis of upper limb and mononeuritis multiplex | 1.60 (1.25, 2.04)** | 106 | 810 |
| 355 | Mononeuritis of lower limb | 1.05 (0.82, 1.34) | 101 | 1561 |
| 360 | Disorders of the globe | 1.19 (0.58, 2.44) | 12 | 152 |
| 367 | Disorders of refraction and accommodation | 1.15 (1.00, 1.32) | 321 | 4465 |
| 368 | Visual disturbances | 1.64 (1.19, 2.26)* | 57 | 462 |
| 370 | Keratitis | 1.01 (0.58, 1.77) | 15 | 215 |
| 371 | Corneal opacity and other disorders of cornea | 1.02 (0.59, 1.76) | 19 | 285 |
| 372 | Disorders of conjunctiva | 0.95 (0.84, 1.06) | 462 | 7420 |
| 373 | Inflammation of eyelids | 0.79 (0.63, 0.99) | 103 | 1870 |
| 374 | Other disorders of eyelids | 0.90 (0.55, 1.48) | 22 | 363 |
| 375 | Disorders of lacrimal system | 1.50 (1.15, 1.97)* | 81 | 722 |
| 378 | Strabismus and other disorders of binocular eye movements | 1.67 (1.06, 2.61) | 30 | 289 |
| 379 | Other disorders of eye | 1.52 (1.18, 1.97)* | 90 | 916 |
| 380 | Disorders of external ear | 1.06 (0.91, 1.22) | 294 | 4553 |
| 381 | Nonsuppurative otitis media and Eustachian tube disorders | 1.17 (0.98, 1.39) | 196 | 2366 |
| 382 | Suppurative and unspecified otitis media | 1.12 (0.92, 1.36) | 146 | 1991 |
| 384 | Other disorders of tympanic membrane | 0.81 (0.45, 1.45) | 14 | 298 |
| 386 | Vertiginous syndromes and other disorders of vestibular system | 2.18 (1.63, 2.92)*** | 75 | 430 |
| 388 | Other disorders of ear | 1.26 (1.07, 1.48)* | 226 | 2587 |
| 389 | Hearing loss | 1.51 (1.17, 1.97)* | 84 | 739 |
| 401 | Essential hypertension | 1.39 (1.08, 1.77)* | 102 | 817 |
| 424 | Other diseases of endocardium | 1.88 (1.21, 2.93)* | 32 | 208 |
| 427 | Cardiac dysrhythmias | 2.81 (1.97, 4.02)*** | 48 | 237 |
| 448 | Disease of capillaries | 1.20 (0.88, 1.64) | 55 | 716 |
| 451 | Phlebitis and thrombophlebitis | 0.56 (0.34, 0.92) | 24 | 478 |
| 454 | Varicose veins of lower extremities | 0.84 (0.70, 1.00) | 201 | 3371 |
| 455 | Hemorrhoids | 1.12 (0.96, 1.31) | 242 | 3172 |
| 458 | Hypotension | 1.12 (0.83, 1.51) | 63 | 721 |
| 459 | Other disorders of circulatory system | 0.69 (0.40, 1.16) | 20 | 433 |
| 460 | Acute nasopharyngitis [common cold] | 1.17 (1.01, 1.35) | 280 | 4021 |
| 461 | Acute sinusitis | 1.07 (0.96, 1.18) | 672 | 9337 |
| 462 | Acute pharyngitis | 0.93 (0.84, 1.03) | 804 | 13225 |
| 463 | Acute tonsillitis | 0.82 (0.75, 0.90)*** | 835 | 15638 |
| 464 | Acute laryngitis and tracheitis | 1.02 (0.85, 1.22) | 191 | 2628 |
| 465 | Acute upper respiratory infections of multiple or unspecified sites | 1.02 (0.94, 1.11) | 1530 | 24429 |
| 466 | Acute bronchitis and bronchiolitis | 0.92 (0.81, 1.03) | 458 | 6852 |
| 470 | Deviated nasal septum | 1.51 (0.96, 2.36) | 26 | 250 |
| 472 | Chronic pharyngitis and nasopharyngitis | 1.12 (0.92, 1.37) | 143 | 1560 |
| 473 | Chronic sinusitis | 1.32 (0.95, 1.82) | 54 | 475 |
| 474 | Chronic disease of tonsils and adenoids | 0.84 (0.55, 1.28) | 33 | 547 |
| 476 | Chronic laryngitis and laryngotracheitis | 1.21 (0.63, 2.32) | 13 | 103 |
| 477 | Allergic rhinitis | 0.94 (0.82, 1.09) | 304 | 4037 |
| 478 | Other diseases of upper respiratory tract | 1.13 (0.98, 1.30) | 298 | 4189 |
| 483 | Pneumonia due to other specified organism | 1.12 (0.67, 1.89) | 20 | 256 |
| 485 | Bronchopneumonia, organism unspecified | 0.53 (0.27, 1.04) | 10 | 240 |
| 486 | Pneumonia, organism unspecified | 1.03 (0.75, 1.40) | 62 | 928 |
| 487 | Influenza | 0.85 (0.70, 1.02) | 168 | 2948 |
| 490 | Bronchitis, not specified as acute or chronic | 1.74 (1.16, 2.60)* | 37 | 361 |
| 493 | Asthma | 1.01 (0.83, 1.22) | 160 | 2137 |
| 521 | Diseases of hard tissues of teeth | 1.33 (1.03, 1.70) | 98 | 978 |
| 522 | Diseases of pulp and periapical tissues | 1.17 (0.96, 1.43) | 155 | 1845 |
| 523 | Gingival and periodontal diseases | 1.30 (0.99, 1.72) | 80 | 836 |
| 524 | Dentofacial anomalies, including malocclusion | 1.56 (1.19, 2.05)* | 78 | 692 |
| 525 | Other diseases and conditions of the teeth and supporting structures | 1.36 (0.96, 1.94) | 44 | 585 |
| 527 | Diseases of the salivary glands | 1.20 (0.58, 2.46) | 10 | 135 |
| 528 | Diseases of the oral soft tissues, excluding lesions specific for gingiva and tongue | 1.08 (0.91, 1.29) | 195 | 2542 |
| 529 | Diseases and other conditions of the tongue | 1.87 (1.12, 3.12) | 24 | 198 |
| 530 | Diseases of esophagus | 1.66 (1.34, 2.06)*** | 134 | 914 |
| 532 | Duodenal ulcer | 1.88 (1.21, 2.92)* | 28 | 162 |
| 533 | Peptic ulcer, site unspecified | 1.43 (0.82, 2.51) | 18 | 127 |
| 535 | Gastritis and duodenitis | 1.62 (1.36, 1.92)*** | 211 | 1633 |
| 536 | Disorders of function of stomach | 1.46 (1.12, 1.92)* | 87 | 681 |
| 540 | Acute appendicitis | 1.06 (0.61, 1.85) | 18 | 198 |
| 550 | Inguinal hernia | 0.98 (0.57, 1.68) | 17 | 267 |
| 553 | Other hernia of abdominal cavity without mention of obstruction or gangrene | 1.32 (0.94, 1.86) | 48 | 520 |
| 558 | Other and unspecified noninfectious gastroenteritis and colitis | 1.11 (0.98, 1.27) | 369 | 4468 |
| 564 | Functional digestive disorders, not elsewhere classified | 1.41 (1.21, 1.65)*** | 256 | 2339 |
| 565 | Anal fissure and fistula | 1.34 (1.00, 1.81) | 65 | 738 |
| 569 | Other disorders of intestine | 0.95 (0.68, 1.35) | 49 | 592 |
| 571 | Chronic liver disease and cirrhosis | 1.38 (0.67, 2.85) | 12 | 76 |
| 574 | Cholelithiasis | 0.97 (0.64, 1.46) | 37 | 457 |
| 575 | Other disorders of gallbladder | 1.16 (0.57, 2.34) | 10 | 131 |
| 579 | Intestinal malabsorption | 1.15 (0.59, 2.24) | 10 | 81 |
| 591 | Hydronephrosis | 1.22 (0.66, 2.26) | 15 | 157 |
| 592 | Calculus of kidney and ureter | 1.45 (0.95, 2.20) | 35 | 238 |
| 595 | Cystitis | 1.08 (0.96, 1.23) | 416 | 5773 |
| 599 | Other disorders of urethra and urinary tract | 1.06 (0.96, 1.19) | 585 | 7755 |
| 610 | Benign mammary dysplasias | 1.11 (0.88, 1.41) | 98 | 1112 |
| 611 | Other disorders of breast | 0.97 (0.83, 1.13) | 249 | 4268 |
| 614 | Inflammatory disease of ovary, fallopian tube, pelvic cellular tissue, and peritoneum | 1.49 (1.16, 1.90)* | 96 | 788 |
| 616 | Inflammatory disease of cervix, vagina, and vulva | 1.05 (0.93, 1.17) | 519 | 7386 |
| 618 | Genital prolapse | 0.97 (0.48, 1.97) | 10 | 112 |
| 620 | Noninflammatory disorders of ovary, fallopian tube, and broad ligament | 1.14 (0.87, 1.48) | 74 | 852 |
| 621 | Disorders of uterus, not elsewhere classified | 0.93 (0.45, 1.92) | 11 | 141 |
| 622 | Noninflammatory disorders of cervix | 0.96 (0.74, 1.24) | 81 | 1081 |
| 623 | Noninflammatory disorders of vagina | 0.96 (0.84, 1.09) | 336 | 4622 |
| 625 | Pain and other symptoms associated with female genital organs | 0.97 (0.79, 1.20) | 134 | 1657 |
| 626 | Disorders of menstruation and other abnormal bleeding from female genital tract | 1.10 (1.00, 1.20) | 940 | 13828 |
| 627 | Menopausal and postmenopausal disorders | 1.24 (0.71, 2.18) | 18 | 191 |
| 628 | Infertility, female | 0.81 (0.72, 0.92)* | 373 | 6049 |
| 629 | Other disorders of female genital organs | 0.93 (0.58, 1.49) | 26 | 293 |
| 632 | Missed abortion | 0.84 (0.70, 1.01) | 179 | 3073 |
| 633 | Ectopic pregnancy | 0.95 (0.67, 1.33) | 52 | 715 |
| 634 | Spontaneous abortion | 0.90 (0.74, 1.10) | 138 | 2041 |
| 635 | Legally induced abortion | 0.68 (0.34, 1.34) | 11 | 164 |
| 640 | Hemorrhage in early pregnancy | 1.06 (0.92, 1.24) | 270 | 3858 |
| 641 | Antepartum hemorrhage, abruptio placentae, and placenta previa | 0.37 (0.17, 0.80) | 10 | 331 |
| 642 | Hypertension complicating pregnancy, childbirth, and the puerperium | 1.12 (0.89, 1.40) | 116 | 1371 |
| 643 | Excessive vomiting in pregnancy | 1.16 (0.96, 1.41) | 166 | 1842 |
| 644 | Early or threatened labor | 1.07 (0.89, 1.28) | 182 | 2285 |
| 645 | Late pregnancy | 0.95 (0.65, 1.38) | 37 | 798 |
| 646 | Other complications of pregnancy, not elsewhere classified | 0.98 (0.84, 1.15) | 237 | 3464 |
| 648 | Other current conditions in the mother classifiable elsewhere, but complicating pregnancy, childbirth, or the puerperium | 0.95 (0.85, 1.06) | 619 | 9070 |
| 650 | Normal delivery | 0.58 (0.33, 1.02) | 16 | 525 |
| 651 | Multiple gestation | 0.89 (0.69, 1.15) | 82 | 1152 |
| 652 | Malposition and malpresentation of fetus | 0.81 (0.50, 1.32) | 21 | 363 |
| 654 | Abnormality of organs and soft tissues of pelvis | 0.94 (0.72, 1.24) | 80 | 1619 |
| 655 | Known or suspected fetal abnormality affecting management of mother | 1.33 (0.71, 2.49) | 14 | 129 |
| 656 | Other known or suspected fetal and placental problems affecting management of mother | 1.30 (0.90, 1.88) | 39 | 512 |
| 657 | Polyhydramnios | 1.00 (0.66, 1.52) | 32 | 508 |
| 658 | Other problems associated with amniotic cavity and membranes | 1.00 (0.68, 1.47) | 34 | 615 |
| 659 | Other indications for care or intervention related to labor and delivery, not elsewhere classified | 0.79 (0.53, 1.20) | 37 | 633 |
| 663 | Umbilical cord complications | 0.69 (0.46, 1.04) | 33 | 767 |
| 669 | Other complications of labor and delivery, not elsewhere classified | 0.79 (0.53, 1.17) | 34 | 574 |
| 671 | Venous complications in pregnancy and the puerperium | 0.75 (0.62, 0.89)* | 192 | 3846 |
| 675 | Infections of the breast and nipple associated with childbirth | 0.52 (0.29, 0.92) | 14 | 423 |
| 676 | Other disorders of the breast associated with childbirth and disorders of lactation | 1.28 (0.89, 1.84) | 43 | 564 |
| 680 | Carbuncle and furuncle | 1.05 (0.77, 1.44) | 55 | 704 |
| 681 | Cellulitis and abscess of finger and toe | 1.13 (0.88, 1.47) | 90 | 1309 |
| 682 | Other cellulitis and abscess | 1.06 (0.88, 1.29) | 152 | 2103 |
| 683 | Acute lymphadenitis | 0.95 (0.60, 1.51) | 24 | 395 |
| 684 | Impetigo | 1.06 (0.69, 1.63) | 32 | 467 |
| 686 | Other local infections of skin and subcutaneous tissue | 1.04 (0.79, 1.37) | 76 | 1036 |
| 690 | Erythematosquamous dermatosis | 1.21 (0.91, 1.60) | 70 | 879 |
| 691 | Atopic dermatitis and related conditions | 0.85 (0.66, 1.09) | 93 | 1665 |
| 692 | Contact dermatitis and other eczema | 0.92 (0.81, 1.03) | 448 | 7426 |
| 695 | Erythematous conditions | 1.10 (0.80, 1.51) | 53 | 736 |
| 696 | Psoriasis and similar disorders | 0.79 (0.58, 1.08) | 60 | 1112 |
| 698 | Pruritus and related conditions | 1.10 (0.92, 1.30) | 192 | 2382 |
| 700 | Corns and callosities | 0.87 (0.62, 1.22) | 56 | 971 |
| 701 | Other hypertrophic and atrophic conditions of skin | 1.26 (1.00, 1.59) | 109 | 1273 |
| 702 | Other dermatoses | 0.70 (0.43, 1.16) | 20 | 424 |
| 703 | Diseases of nail | 1.12 (0.87, 1.43) | 99 | 1596 |
| 704 | Diseases of hair and hair follicles | 1.29 (1.08, 1.55)* | 180 | 1960 |
| 705 | Disorders of sweat glands | 1.05 (0.71, 1.53) | 37 | 577 |
| 706 | Diseases of sebaceous glands | 1.22 (1.07, 1.39)* | 366 | 4945 |
| 708 | Urticaria | 0.92 (0.74, 1.15) | 117 | 1691 |
| 709 | Other disorders of skin and subcutaneous tissue | 0.94 (0.75, 1.16) | 118 | 1663 |
| 710 | Diffuse diseases of connective tissue | 1.07 (0.55, 2.06) | 11 | 149 |
| 713 | Arthropathy associated with other disorders classified elsewhere | 2.01 (1.16, 3.49) | 20 | 144 |
| 714 | Rheumatoid arthritis and other inflammatory polyarthropathies | 1.36 (0.74, 2.50) | 18 | 138 |
| 715 | Osteoarthrosis and allied disorders | 1.20 (0.64, 2.26) | 15 | 138 |
| 716 | Other and unspecified arthropathies | 1.06 (0.56, 2.01) | 14 | 167 |
| 717 | Internal derangement of knee | 1.25 (0.86, 1.80) | 43 | 494 |
| 719 | Other and unspecified disorders of joint | 1.31 (1.15, 1.50)*** | 355 | 3612 |
| 720 | Ankylosing spondylitis and other inflammatory spondylopathies | 1.98 (0.97, 4.02) | 11 | 77 |
| 721 | Spondylosis and allied disorders | 1.25 (0.65, 2.39) | 13 | 98 |
| 722 | Intervertebral disc disorders | 1.52 (1.06, 2.18) | 42 | 304 |
| 723 | Other disorders of cervical region | 1.70 (1.48, 1.95)*** | 344 | 2627 |
| 724 | Other and unspecified disorders of back | 1.37 (1.25, 1.50)*** | 934 | 10222 |
| 726 | Peripheral enthesopathies and allied syndromes | 1.03 (0.86, 1.23) | 190 | 2425 |
| 727 | Other disorders of synovium, tendon, and bursa | 1.06 (0.82, 1.35) | 94 | 1186 |
| 728 | Disorders of muscle, ligament, and fascia | 1.37 (1.05, 1.81) | 79 | 698 |
| 729 | Other disorders of soft tissues | 1.28 (1.14, 1.43)*** | 519 | 5500 |
| 733 | Other disorders of bone and cartilage | 1.86 (1.28, 2.71)* | 48 | 319 |
| 734 | Flat foot | 0.93 (0.65, 1.34) | 45 | 642 |
| 736 | Other acquired deformities of limbs | 1.85 (0.96, 3.56) | 13 | 100 |
| 737 | Curvature of spine | 1.53 (0.94, 2.50) | 23 | 252 |
| 754 | Certain congenital musculoskeletal deformities | 1.26 (0.69, 2.29) | 15 | 184 |
| 757 | Congenital anomalies of the integument | 1.38 (0.90, 2.14) | 27 | 309 |
| 764 | Slow fetal growth and fetal malnutrition | 1.02 (0.72, 1.46) | 41 | 791 |
| 766 | Disorders relating to long gestation and high birthweight | 0.90 (0.45, 1.78) | 11 | 212 |
| 780 | General symptoms | 1.60 (1.47, 1.75)*** | 1282 | 13993 |
| 781 | Symptoms involving nervous and musculoskeletal systems | 3.08 (1.93, 4.91)*** | 25 | 125 |
| 782 | Symptoms involving skin and other integumentary tissue | 1.18 (1.02, 1.38) | 279 | 3653 |
| 783 | Symptoms concerning nutrition, metabolism, and development | 1.98 (1.58, 2.47)*** | 123 | 850 |
| 784 | Symptoms involving head and neck | 1.61 (1.47, 1.76)*** | 1116 | 11125 |
| 785 | Symptoms involving cardiovascular system | 2.09 (1.83, 2.40)*** | 373 | 2360 |
| 786 | Symptoms involving respiratory system and other chest symptoms | 1.28 (1.16, 1.41)*** | 846 | 9617 |
| 787 | Symptoms involving digestive system | 1.48 (1.32, 1.67)*** | 491 | 4503 |
| 788 | Symptoms involving urinary system | 1.13 (0.97, 1.31) | 283 | 3097 |
| 789 | Other symptoms involving abdomen and pelvis | 1.23 (1.12, 1.34)*** | 1277 | 16281 |
| 790 | Nonspecific findings on examination of blood | 1.06 (0.85, 1.32) | 125 | 1473 |
| 791 | Nonspecific findings on examination of urine | 0.68 (0.40, 1.14) | 19 | 308 |
| 794 | Nonspecific abnormal results of function studies | 0.67 (0.35, 1.27) | 13 | 234 |
| 795 | Other and nonspecific abnormal cytological, histological, immunological and DNA test findings | 0.60 (0.29, 1.25) | 11 | 202 |
| 799 | Other ill-defined and unknown causes of morbidity and mortality | 1.31 (1.18, 1.46)*** | 702 | 8620 |
| 813 | Fracture of radius and ulna | 0.83 (0.44, 1.58) | 12 | 141 |
| 816 | Fracture of one or more phalanges of hand | 1.03 (0.54, 1.97) | 11 | 127 |
| 824 | Fracture of ankle | 1.62 (0.82, 3.20) | 12 | 128 |
| 825 | Fracture of one or more tarsal and metatarsal bones | 1.29 (0.70, 2.36) | 13 | 176 |
| 845 | Sprains and strains of ankle and foot | 1.02 (0.80, 1.31) | 94 | 1418 |
| 847 | Sprains and strains of other and unspecified parts of back | 1.49 (1.18, 1.88)** | 101 | 763 |
| 854 | Intracranial injury of other and unspecified nature | 3.04 (1.75, 5.26)*** | 20 | 83 |
| 873 | Other open wound of head | 0.76 (0.38, 1.51) | 10 | 153 |
| 879 | Open wound of other and unspecified sites, except limbs | 0.97 (0.46, 2.04) | 11 | 172 |
| 883 | Open wound of finger(s) | 0.91 (0.61, 1.37) | 31 | 491 |
| 910 | Superficial injury of face, neck, and scalp except eye | 1.19 (0.86, 1.64) | 58 | 741 |
| 918 | Superficial injury of eye and adnexa | 1.16 (0.65, 2.09) | 14 | 239 |
| 919 | Superficial injury of other, multiple, and unspecified sites | 1.18 (0.80, 1.74) | 38 | 512 |
| 920 | Contusion of face, scalp, and neck except eye(s) | 1.44 (0.91, 2.27) | 26 | 229 |
| 922 | Contusion of trunk | 1.40 (1.09, 1.78)* | 99 | 822 |
| 923 | Contusion of upper limb | 1.12 (0.86, 1.44) | 88 | 968 |
| 924 | Contusion of lower limb and of other and unspecified sites | 1.24 (1.02, 1.52) | 145 | 1606 |
| 945 | Burn of lower limb(s) | 2.04 (1.04, 3.98) | 14 | 109 |
| 949 | Burn, unspecified | 1.09 (0.76, 1.54) | 43 | 536 |
| 958 | Certain early complications of trauma | 0.93 (0.59, 1.44) | 32 | 516 |
| 959 | Injury, other and unspecified | 1.05 (0.67, 1.66) | 27 | 371 |
| 989 | Toxic effect of other substances, chiefly nonmedicinal as to source | 1.26 (0.63, 2.51) | 10 | 149 |
| 994 | Effects of other external causes | 3.91 (1.82, 8.42)** | 13 | 48 |
| 995 | Certain adverse effects not elsewhere classified | 0.92 (0.76, 1.11) | 162 | 2216 |
| E81 | Motor vehicle traffic accidents | 1.54 (1.16, 2.05)* | 70 | 514 |
| E92 | Late effects of accidental injury | 1.47 (1.01, 2.13) | 42 | 397 |
| E96 | Homicide and injury purposely inflicted by other persons | 1.13 (0.76, 1.68) | 34 | 409 |

^a^ Odds ratios are adjusted for SES, maternal age at delivery, total number of diagnoses during 21 months period before delivery, and year of delivery

*: Q-value <0.05; **: Q-value <0.01; ***: Q-value <0.001

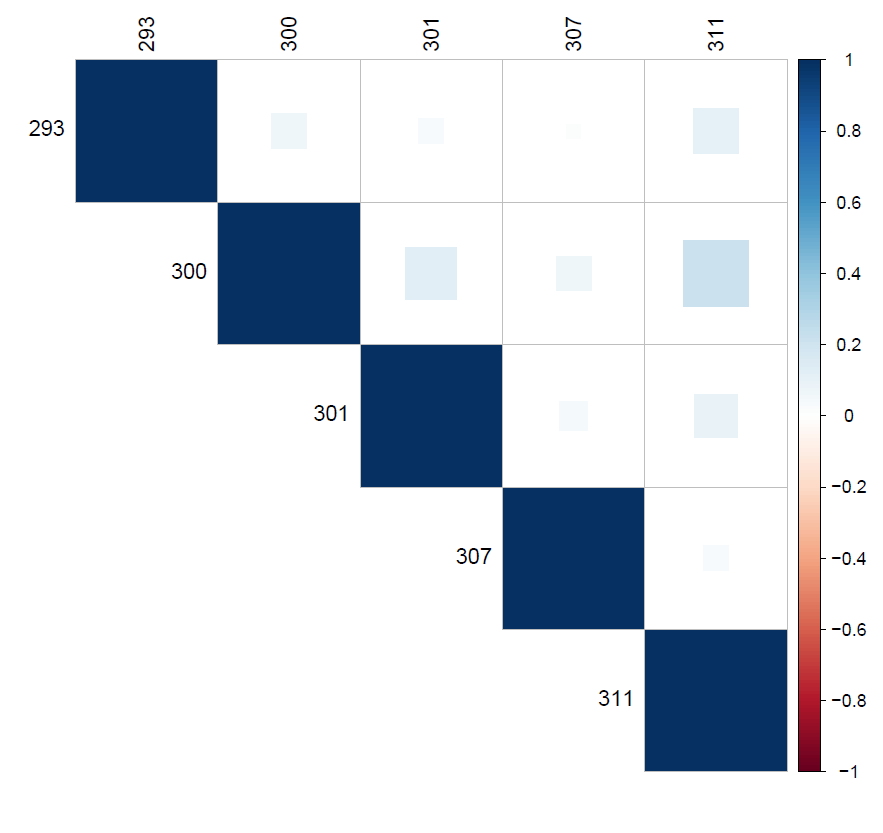

**Figure S3.** Correlation between maternal psychiatric diagnoses evaluated for their association with maternal diagnoses around pregnancy period.
Note: All correlations have a p-value of < 0.05.

**Table S2.** Comparison of results obtained using (1) logistic regression (no imputation) and (2) logistic regression with imputation, for the associations between maternal ICD-9 level 1 diagnostic categories and ICD-9 level 1 mental health diagnosis

|  | **Multivariable logistic regression model^a^ no imputation (N=76,783)** | **Multivariable logistic regression model^a^ with imputation (N=84,740)** |
| --- | --- | --- |
| **Level 1 ICD-9 diagnostic category** | **OR (95%CI)** | **OR (95%CI)** |
| **001-139:** Infectious and Parasitic Diseases | 1.09 (1.00,1.19)† | 1.07 (0.99, 1.16) |
| **140-239:** Neoplasms | 1.06 (0.92,1.21) | 1.06 (0.93, 1.20) |
| **240-279:** Endocrine, Nutritional and Metabolic Diseases, and Immunity Disorders | 1.12 (1.02,1.24)† | 1.14 (1.04, 1.25)†† |
| **280-289:** Diseases of Blood and Blood-Forming Organs | 1.17 (1.05,1.29)†† | 1.16 (1.05, 1.28)†† |
| **320-389:** Diseases of the Nervous System and Sense Organs | 1.25 (1.15,1.36)††† | 1.26 (1.17, 1.37)††† |
| **390-459:** Diseases of the Circulatory System | 1.13 (1.02,1.25)† | 1.15 (1.04, 1.27)†† |
| **460-519:** Diseases of the Respiratory System | 0.95 (0.85,1.06) | 0.98 (0.88, 1.09) |
| **520-579:** Diseases of the Digestive System | 1.25 (1.15,1.37)††† | 1.24 (1.14, 1.35)††† |
| **580-629:** Diseases of the Genitourinary System | 1.05 (0.96,1.15) | 1.05 (0.96, 1.14) |
| **630-677:** Complications of Pregnancy, Childbirth, and the Puerperium | 0.95 (0.87,1.03) | 0.95 (0.87, 1.03) |
| **680-709:** Diseases of the Skin and Subcutaneous Tissue | 1.05 (0.96,1.14) | 1.03 (0.95, 1.11) |
| **710-739:** Diseases of the Musculoskeletal System and Connective Tissue | 1.33 (1.22,1.45)††† | 1.34 (1.23, 1.45)††† |
| **740-759:** Congenital Anomalies | 1.16 (0.90,1.49) | 1.10 (0.86, 1.40) |
| **760-779:** Certain Conditions Originating in the Perinatal Period | 1.03 (0.79,1.36) | 1.09 (0.84, 1.40) |
| **780-799:** Symptoms, Signs, and Ill-Defined Conditions | 1.69 (1.50,1.89)††† | 1.68 (1.51, 1.88)††† |
| **800-999:** Injury and Poisoning | 1.10 (1.00,1.21)† | 1.11 (1.01, 1.21)† |
| **E800-E999:** Supplementary Classification of External Causes of Injury and Poisoning | 1.39 (1.13,1.71)†† | 1.42 (1.16, 1.73)††† |
| ^a^ model adjusted for SES, maternal age at delivery, total number of diagnoses during 21 months period before delivery, year of delivery, and all the ICD-9 level 1 diagnostic categories presented in this table  † : P-value <0.05; †† : P-value <0.01; ††† :P-value <0.001 | | |
